# Single-nuclei paired multiomic analysis of young, aged, and Parkinson’s disease human midbrain reveals age- and disease-associated glial changes and their contribution to Parkinson’s disease

**DOI:** 10.1101/2022.01.18.22269350

**Authors:** Levi Adams, Min Kyung Song, Yoshiaki Tanaka, Yoon-Seong Kim

**Author notes:** These two authors contributed equally (Levi Adams, Min Kyung Song). Corresponding authors: Yoon-Seong Kim, M.D., Ph.D., RWJMS-Institute for Neurological Therapeutics at Rutgers 683 Hoes Lane West, Piscataway, NJ 08854; Yoshiaki Tanaka, Ph.D., Maisonneuve-Rosemont Hospital Research Center (CRHMR), Department of Medicine, University of Montreal, QC, Canada, 5415 boulevard de I’Assomption, Montreal, Quebec, H1T 2M4, Canada.

## Abstract

Age is the primary risk factor for Parkinson’s disease (PD), but how aging changes the expression and regulatory landscape of the brain remains unclear. Here, we present a single-nuclei multiomic study profiling shared gene expression and chromatin accessibility of young, aged and PD post-mortem midbrain samples. Combined multiomic analysis along a pseudopathogenesis trajectory reveals all glial cell types are affected by age, but microglia and oligodendrocytes are further altered in PD. We present evidence for a novel disease-associated oligodendrocyte subtype and identify genes lost over the aging and disease process, including CARNS1, that may predispose healthy cells to develop a disease-associated phenotype. Peak-gene association analysis from paired data identifies 89 PD-associated SNP loci, including five in MAPT, that show differential association with gene expression in disease-associated oligodendrocytes. Our study suggests a previously undescribed role for oligodendrocytes in aging and PD pathogenesis.

---

Parkinson’s disease (PD) is the second most common neurodegenerative disease and is estimated to affect more than ten million people globally^1^. PD prevalence increases from 41 cases per 100,000 individuals in the fourth decade of life to 425 per 100,000 in the sixth decade of life and to 1908 per 100,000 in the eighth decade of life^2^. During PD pathogenesis, dopaminergic neurons in the substantia nigra degenerate, resulting in neurological, cognitive and motor symptoms^3^. While many genetic components and environmental risk factors have been identified, the primary risk factor for PD is aging^4^.

Despite the connection between aging and PD, relatively little attention has been given to the changes in the aging midbrain and how these changes may predispose individuals to development of PD. Transcriptomic studies of mouse or fly brains have provided some insight into aging, but have not focused on the midbrain^5, 6^. Glaab and Schneider^7^ used microarray datasets to investigate shared pathways and network alterations in aging and PD, but there remains a lack of information on how alterations in gene expression during aging affect the different cell types in the midbrain and how they contribute to PD pathogenesis.

The field of single-cell transcriptomics and epigenomics has facilitated important advances in understanding how gene expression and chromatin accessibility contribute to neurodegeneration^8–10^. Recent single-cell expression studies of the midbrain have highlighted a potential role of oligodendrocytes in the midbrain in PD pathogenesis, supported by strong genome-wide association study (GWAS) and transcriptomic data^11–14^. This is a surprising finding because vulnerable dopaminergic neurons that are lost during PD pathogenesis are sparsely myelinated^15^. Despite these crucial insights, there remains a lack of cell type-specific high-resolution data on what distinguishes ‘healthy’ aging from neurodegeneration and an incomplete picture of how the epigenetic landscape changes during aging and PD.

To address this gap, we isolated nuclei from the substantia nigra of post-mortem midbrains of young and aged donors with no neurological disease as well as PD patients and compared the single-nuclei transcriptome and genome-wide chromatin accessibility simultaneously from each nucleus along aging and PD trajectories. This approach allows us to directly infer *cis-*acting elements that contribute to gene expression in the same cells and provides a more complete picture of how the aging process affects gene regulation and expression in distinct cell types in the midbrain. This multiomic analysis reveals a novel disease-associated oligodendrocyte subset that may contribute to PD pathogenesis.

## Results

### Simultaneous single-nuclei RNA-seq and ATAC-seq profiling of the human post-mortem midbrain

We obtained frozen, post-mortem midbrain samples of young (mean 24 years old) and aged (mean 75 years old) neurologically healthy donors and patients diagnosed with PD (mean 81 years old) (Supplementary Table 1). We isolated nuclei from the substantia nigra and performed paired single-nuclei RNA-sequencing (snRNA-seq) and single-nuclei Assay for Transposase Accessible Chromatin sequencing (snATAC-seq) on each nucleus (10X Genomics Single Cell Multiome ATAC + Gene Expression kit) (Fig. 1a, Supplementary Fig. 1a-c). After filtering to remove low-quality reads or potential multiplets (Methods), we retained 69,289 high-quality nuclei from 31 individuals (9 young donors, 8 aged donors, 14 PD patients), showing low rates of doublets based on gene expression and robust ATAC data quality (Supplementary Fig. 1d-g). Using these nuclei, we performed batch correction, variable gene and principal component analysis and UMAP dimensional reduction with Seurat v4^16^ (Methods). We identified 23 separate clusters of nuclei in our snATAC-seq and snRNA-seq datasets (Extended Data Fig. 1a). Interrogation of gene expression patterns of known cell type markers^10, 17, 18^ allowed us to classify our nuclei into seven major cell types (Fig. 1b,d, Extended Data Fig. 1b): Neurons (N), Oligodendrocytes (ODC), Astrocytes (AS), Microglia (MG), Oligodendrocyte Precursor Cells (OPC), Endothelial Cells (EC), and peripheral immune cells/T-cells (T). As expected, clustering based on ATAC profiling generated the same major clustering patterns for these cells with distinctive chromatin accessibility profiles at key loci (Fig. 1c,e, Extended Data Fig. 1b). We found the vast majority of cells in the midbrain are ODC, followed by MG, OPC and AS (Fig. 1f, Supplementary Fig. 1h). Similar to recently published data^14^, we did note a statistical difference in MG and ODC populations between groups, with a significantly higher proportion of ODC (86.9%) and lower proportion of MG (3.8%) in the aged group compared to either the young (ODC, 76.5%; MG, 9.7%) or PD group (ODC, 74.0%; MG, 9.4%) (Supplementary Table 2).

**Fig. 1.**
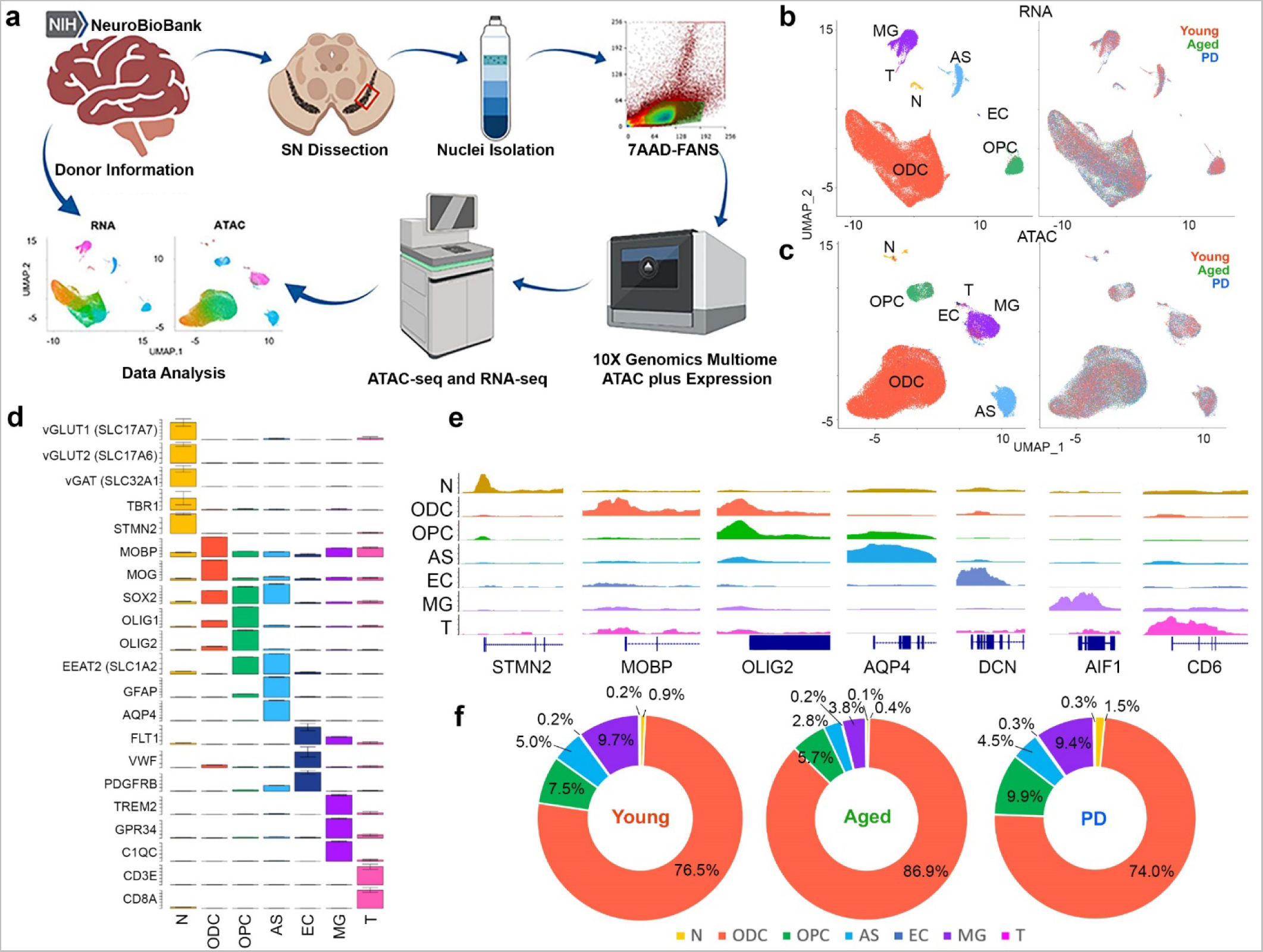
Multiomic analysis of human midbrain. **a**, Schematic of isolation and single-nuclei ATAC plus Gene Expression analysis for human post-mortem midbrain. **b,c,** UMAP visualization of single nuclei by RNA **(b)** and ATAC **(c)** profiles. Nuclei are colored by identified cell type (left) and donor type (right). **d,** Expression of cell type-specific genes in each annotated cell type cluster. **e,** Chromatin accessibility at cell type-specific genes in each annotated cell type cluster. **f,** Percentage of each cell type passing QC criteria identified in young, aged and PD. Student’s *t*-test shows ODC are significantly changed from young to aged (p-value, 0.002), and aged to PD (p-value, 0.023). MG are significantly changed from young to aged (p-value, 0.007) and aged to PD (p-value, 0.042).

Genes with cell type-specific expression patterns also showed differential ATAC peaks nearest to the transcription start site (TSS), which implies unique promoter accessibility for each cell type (Extended Data Fig. 1c), and distinctive enrichment of cell type-specific transcription factor binding motifs (Extended Data Fig. 1d). Within each cell type, we analyzed differentially expressed genes (DEG) between young and aged groups, as well as aged and PD groups (Supplementary Table 3). Several genes that have been previously associated with neurological disorders, such as NEAT1, FKBP5 and SLC38A2, were differentially expressed in multiple cell types (Supplementary Fig. 2a,b, Methods). We identified cell type-specific changes in gene expression across groups (Supplementary Fig. 2c,d,f) that showed potential alterations in neurological function and metabolic pathways (Supplementary Fig. 2e,g). The majority of differentially expressed genes were cell-type specific (Supplementary Fig. 3a). Integration of chromatin accessibility data showed that the accessibility of the nearest peak to the TSS of differentially expressed genes generally did not change between young and aged or aged and PD (Supplementary Fig. 3b-e) within the same cell type. Only approximately 10% of putative promoters for genes differentially expressed during aging or PD pathogenesis had correlation values over 0.85, as opposed to cell-type specific genes which strongly correlate with promoter peaks (Extended Data Fig. 1c), suggesting that the expression of DEG between groups might be controlled by other distal regulatory elements, not by a promoter.

We noticed an inverse correlation between number of nuclei and number of DEG for each group, with the larger groups having fewer DEG (Fig. 1f, Supplementary Fig. 2c). This is a common effect in transcriptomics known as Simpson’s Paradox (thoroughly discussed by Cole Trapnell^19, 20^). When we examine the gene expression signatures of previously defined functional subtypes^21–26^ (Methods), we saw transcriptionally distinct clusters. We identified clusters corresponding to newly formed ODC, myelin forming ODC and mature ODC (Extended Data Fig. 1e, Supplementary Fig. 4a,d). As previously reported, these clusters showed minimal overlap^23, 26^. We observed clusters with high expression of genes relating to neuronal and synaptic support (such as NRXN3 and NFASC), and clusters with increased expression of ODC-neuron adhesion markers (such as STMN1 and HAPLN2). Microglia signatures including homeostatic MG, aging MG and a small population of Stage1 MG (TREM2-independent) disease-associated MG (Extended Data Fig. 1f, Supplementary Fig. 4b,e)^21, 24^. Astrocytes included signatures for disease-associated astrocytes, A1 reactive astrocytes, and GFAP-low astrocytes (Extended Data Fig. 1g, Supplementary Fig. 4c,f)^22, 25^.

### Multiomic analysis of peak-gene associations

The paired single-nuclei gene expression and ATAC-seq data from the same nuclei provides a unique advantage because it allows us to infer the relationship between chromatin accessibility and gene expression in *cis*. We utilized the analytical framework developed by Ma *et al.* to analyze paired snRNA-seq and snATAC-seq data^8^ to generate peak-gene associations (Fig. 2a). Correlated associations were calculated in the chromatin regions within ±500 kb of the TSS of annotated genes where there is co-variation between chromatin accessibility and gene expression. In each cell type, an average of 193,732 significantly peak-gene associations (5.3 peaks per gene) were identified. Notably, we found most peak-gene associations (66.6-92.2%) were exclusive to young, aged or PD, with a minority (0.274-10.6%) shared between groups (Supplementary Fig. 5a,b). Most shared peak-gene associations (50.9-56.6%) were near the TSS (within 5 kb) and unique peaks were linked in more distal regions, suggesting expression changes between groups are likely driven by more distal regions such as enhancers (Supplementary Fig. 5c). In agreement with previous data^8^, we also observed that not all peaks were connected to a gene (91,177 out of 210,609 peaks were connected to at least one gene), and saw an average of 6,812 genes connecting to more than 10 peaks (Supplementary Fig. 5d-e). Comparing peak-gene associations in ODC between groups identified a subset of genes whose correlated associations were significantly changed during aging and PD progression (Fig. 2b). Increased peak-gene associations in both aged and PD groups compared to young (Fig. 2b, red dots) may indicate aging-specific changes, including the aging marker NEAT1. Conversely, some associations were unchanged between young and aged, but increased only in the PD cohort, including genes previously linked with longevity such as RASGRF1^27^ and MSRA^28^ (Fig. 2b, blue dots). Peak-gene associations for NEAT1 were largely shared in older (aged and PD) groups compared to young samples (Fig. 2c, Extended Data Fig. 2a), while for RASGRF1, similar patterns were observed in normal control donors (young and aged) with much higher associations compared to PD (Fig. 2d, Extended Data Fig. 2b). Analysis of cis-regulatory motifs of the 43 correlated peaks for NEAT1 specific to the older groups (aged and PD) showed enrichment of binding motifs for transcription factors related to aging such as EGR1/2^29^ (Extended Data Fig. 2a, c). For RASGRF1, we found 61 significant shared peaks in the control groups (young and aged), and enriched motifs for NRF2 and ASCL1, which are related to brain health and neurogenesis^30, 31^. These motifs were not utilized in PD samples (Extended Data Fig. 2b, d).

**Fig. 2.**
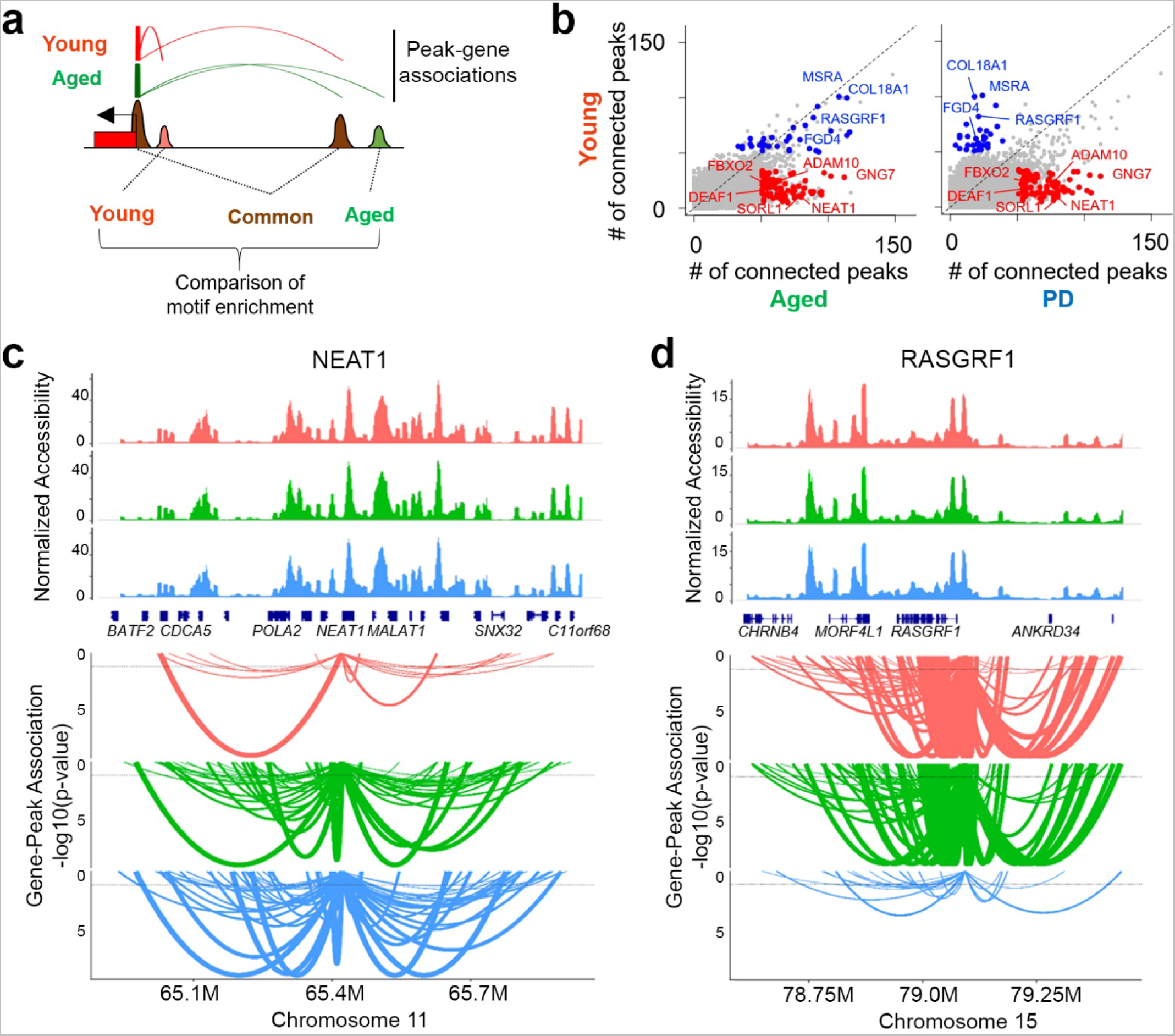
Analysis of gene-peak connections in human midbrain. **a**, Schematic of gene-peak analysis between samples. **b,** Comparison of the number of connected peaks in each gene between young and aged (left), and aged and PD (right) midbrain. Some genes show altered peak-connections from young in both aged and PD (red dots). Some genes show altered peak-connections only in PD (blue dots). **c,d,** Gene-peak connection plots for **(c)** NEAT1 and **(d)** RASGRF1 between young, aged and PD midbrain. Top panel: normalized ATAC read count distribution. Bottom panel: arcs show ATAC peaks significantly correlated with gene expression. Arc height represents statistical significance.

### Defining pseudopathogenesis trajectory in oligodendrocytes

Next, we analyzed each major cell type (ODC, MG, OPC and AS) individually to explore potential changes in gene expression and epigenetic dynamics during the aging and disease process. To highlight heterogeneity and subtle changes within each cell type, we re-clustered each type of cell individually (Methods). Surprisingly, we noticed that for both ATAC and RNA data, nuclei from normal and control groups clustered more closely than PD nuclei (Fig. 3a) although they did not cluster by individual donors (Supplementary Fig. 6a). To identify potential transitions in gene expression and chromatin accessibility between clusters, we performed pseudotime analysis using Monocle3^32^. Pseudotime analysis generated a striking trajectory in both snRNA and snATAC clusters moving from young to aged and then to PD patients in a way we termed pseudopathogenesis (Fig. 3a). Taking advantage of the paired multiomic nature of our data, we combined the snRNA and snATAC pseudopathogenesis scores and corrected for error to establish a combined pseudopathogenesis (cPP) trajectory (Fig. 3b). Comparison of the cPP scores between groups exhibited significant increases from young to aged, and from aged to PD (Fig. 3c). Next, we investigated differentially expressed genes and peaks across the cPP trajectory and identified 299 increased and 474 decreased genes, and 912 increased and 1590 decreased peaks (Fig. 3d, Extended Data Fig. 3a, Supplementary Fig. 6b, Supplementary Table 4). Notably, aging or PD relevant genes that we identified in peak-gene association analysis, such as NEAT1 and RASGRF1 (Fig. 2c,d), were also correlated with cPP (Extended Data Fig. 3b). Gene ontology analysis showed that pathways related to Response to Unfolded Protein, Chaperone-Mediated Autophagy (CMA) and Negative Regulation of Cell Death were increased while Myelination, Receptor Clustering and Regulation of Membrane Potential were decreased with advancing pseudopathogenesis (Fig. 3d). Of note, we observed an increase in RPS family genes that have been previously reported to be increased during aging6 (Supplementary Table 4). We noticed that almost 80% of ODC had low cPP scores of less than 7 (99.8% of young, 90% of aged and 50% of PD) and gene modules for normal functions like myelination and synaptic support showed little change at lower cPP scores. Gene expression module analysis for Myelination and Neuronal and Synaptic Support exhibited decreases at high cPP values, while gene modules for Unfolded Protein Response and CMA increased (Fig. 3e). We also noticed a distinct polarization in expression of several genes including OPALIN and RBFOX1, as previously reported^14^ (Extended Data Fig. 3c). Notably, both RBFOX1 and OPALIN expression levels peaked at lower cPP scores and then were reduced in high cPP, and were lower in PD compared to young donors (Extended Data Fig. 3d,e).

**Fig. 3.**
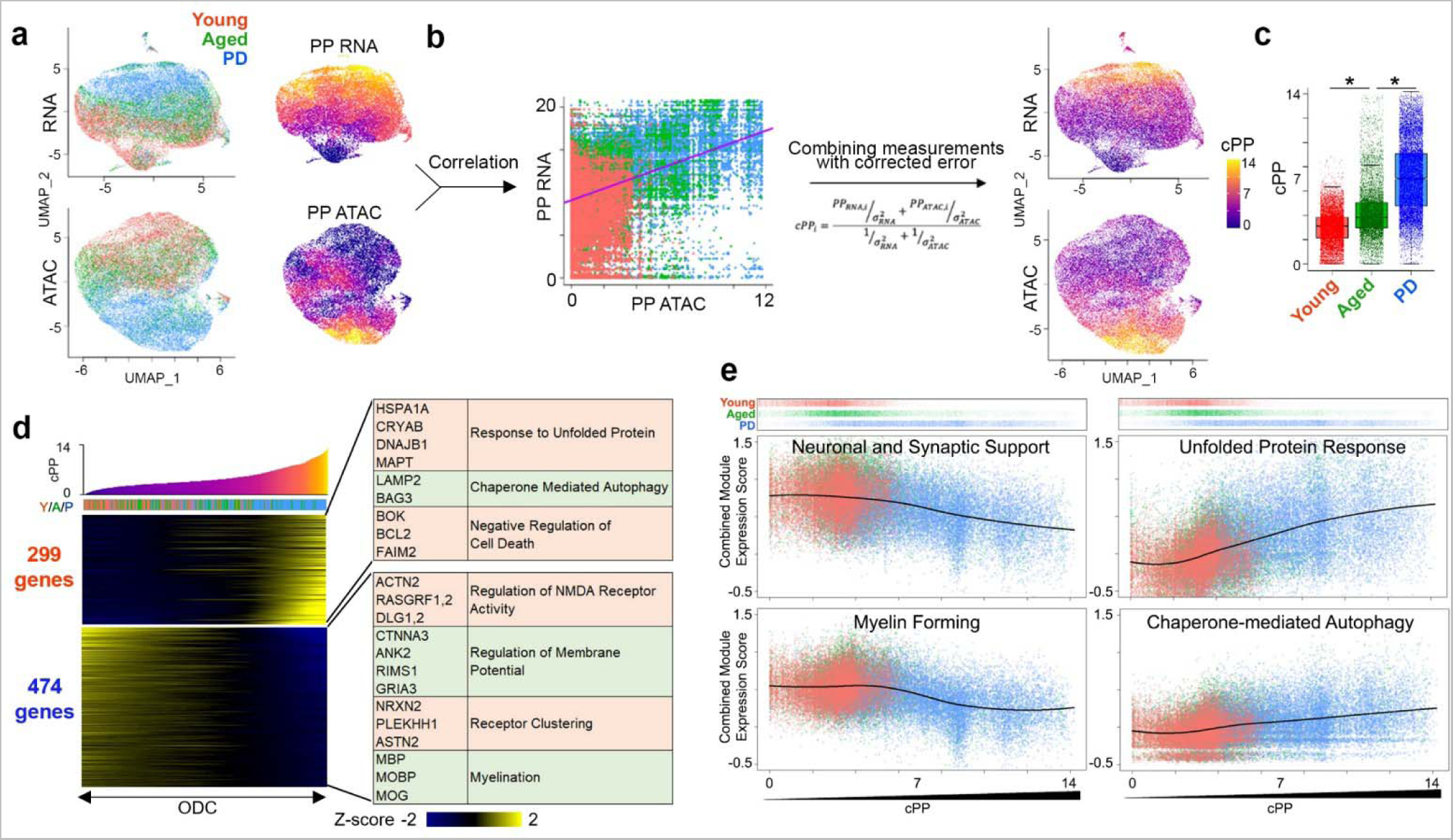
Establishment of pseudopathogenesis trajectory in ODC. **a**, UMAP plot of ODC nuclei colored by young, aged and PD donor (left), and heatmap of RNA- and ATAC-based pseudopathogenesis trajectory (right). **b,** Schematic of calculation of combined pseudopathogenesis (cPP) score from transcriptome and ATAC profiles (Cor = -0.419; p<2.2e-16). **c,** Significant difference in cPP scores of individual ODC nuclei from young, aged and PD midbrain. (One-way ANOVA, p-value for Y/A < 1e- 199, p-value for A/P < 1e-199). **d,** Heatmap showing genes correlated with cPP trajectory. X-axis represents individual cells sorted by cPPs. Y-axis of heatmap represents positively (upper)- and negatively (bottom)-correlated genes. Representative genes and significant GO terms are shown in right panel (Cor > 0.1 or < -0.1). **e,** Change in gene expression modules across cPP trajectory in ODC. Top panel shows individual ODC nuclei along cPP scores and donor group. X-axis shows cPP score. Y-axis is combined expression score for all genes in the expression module.

### Pseudopathogenesis analysis shows distinct changes in microglia

We applied the same analytical framework to microglia and generated a cPP score for each nucleus (Fig. 4a,b, Supplementary Fig. 7a). We found significant increases in cPP between young, aged and PD samples for MG (Fig. 4c). While 894 genes were increased as cPP score increased, 254 genes were decreased (Fig 4d, Supplementary Table 4). Gene ontology analysis of cPP-relevant genes showed a loss of cell adhesion and chemotaxis and elevated immune activation and cytokine-mediated signaling pathway (Fig. 4d). Combined gene module analysis showed a decrease in homeostatic gene signatures and increases in aging and stage1 (TREM2 independent) disease-associated MG (Fig. 4e). We also found distinctive ATAC peak changes with cPP progression (Supplementary Fig. 7b). We analyzed astrocyte clusters by applying the same strategy (Extended Data Fig. 4a,b, Supplementary Fig. 8a). There is a statistically significant increase in cPP score from young to aged samples, but no difference between aged and PD nuclei (Extended Data Fig. 4c). Gene module analysis indicated increases in A1-reactive AS signatures, and disease-associated AS signatures near the middle of the trajectory, with a concomitant reduction of GFAP-low module signatures across cPP, as well in three equal-sized groups based on cPP (Extended Data Fig. 4d). Gene ontology analysis of trajectory-relevant gene expression showed generalized increases in apoptosis resistance and CMA pathways, but a reduction in neuronal support (Supplementary Fig. 8b). OPC exhibited similar results to AS (Supplementary Fig. 9a,b,10a) with a significant increase in cPP scores from young to aged nuclei, but no difference in PD samples (Supplementary Fig. 9c). Gene ontology analysis of trajectory-relevant genes showed a similar result as astrocytes with an increase in general stress-resistance markers during cPP progression, but a reduction in differentiation and synaptic signaling pathways (Supplementary Fig. 10b).

**Fig. 4.**
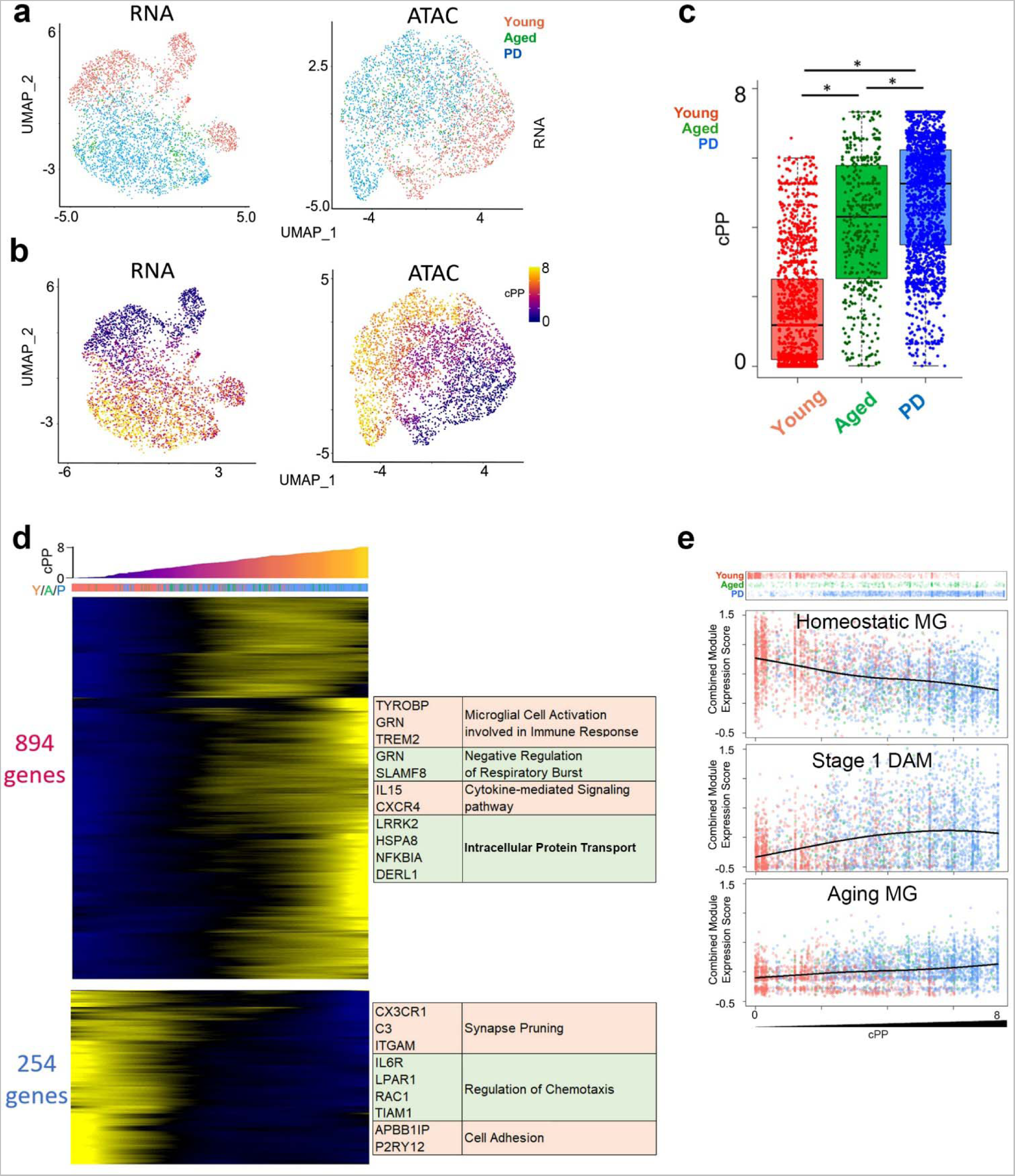
Establishment of cPP trajectory in MG. **a**, UMAP plot of MG nuclei colored by young, aged and PD donors. **b,** UMAP plot of MG nuclei colored by cPP. **c,** Significant difference in cPP scores of individual MG nuclei from young, aged and PD midbrain. (One-way ANOVA, p-value for Y/A < 7e-130, p-value for A/P <9.8e-15). **d,** Heatmap showing genes correlated with cPP trajectory. X-axis represents individual cells sorted by cPP. Y-axis represents positively (upper)- and negatively (bottom)-correlated genes. Representative genes and significant gene ontology terms are shown in right panel (Cor > 0.1 or < -0.1). **e,** Gene expression modules across MG cPP trajectory. Top panel shows individual MG nuclei along cPP scores and donor group. X-axis shows cPP score. Y-axis is combined expression score for all genes in the expression module.

### In-depth analysis of high cPP oligodendrocytes

The psuedopathogenesis trajectory provided a strong tool to analyze the incremental changes as young, healthy cells age and move into a disease-like state. Intriguingly, we noticed that while nearly all cells from young donors had low cPP scores, there was substantial heterogeneity in aged and PD samples (Extended Data Fig. 5a). Plotting cell populations as a function of cPP score revealed three distinct peaks of cells (Fig. 5a), and we identified three populations based on Kernel density estimation^33^. As low-cPP nuclei encapsulated the majority of normal cellular function for oligodendrocytes such as myelination and neuronal support (Extended Data Fig. 3c), we defined this population as ‘Healthy’. The high-cPP population was marked by a loss of canonical ODC functions and increase in stress response genes (Fig. 3e), suggesting they are ‘Disease-associated’ ODC, with an intermediate population we defined as ‘At-Risk’. Healthy cells made up the vast majority of nuclei in our study (76%) and surprisingly 89% of nuclei from aged control donors and 48% of nuclei from PD patients were in the ‘healthy’ population (Fig. 5a, Extended Data Fig. 5b) suggesting that a significant proportion of cells maintain their normal functions even in disease state. We analyzed differentially expressed genes between healthy, at-risk, and disease-associated populations (Supplementary Table 5) and noticed interesting trends when compared to differentially expressed genes between young, aged, and PD cohorts. Some genes were not significantly changed over aging but were changed between healthy and at-risk or disease-associated ODC, suggesting PD-specific changes that are unrelated to the aging process (Fig. 5b, Left). The list includes reductions in multiple genes involved in the myelination pathway (MBP, OPALIN, MOBP) and several glial-neuron adhesion genes (CTNNA2, NRXN2, PLEKHH1), as well as increases in genes not widely explored in PD such as SELENOP, QDPR, SLC38A2, and IGF1R. We also identified a subset of genes that were differentially expressed over aging and further changed toward a disease state (Fig. 5b, Right). These genes are of particular interest as they potentially represent age-related risk factors for the development of PD. They include increases in stress response proteins (HSP90AA1, FKBP5) and well-known neurodegenerative markers such as MAPT. We also observed an age-dependent loss of several genes that are further reduced in disease (RBFOX1, CARNS1). Of particular note, CARNS1, which encodes carnosine synthase1, has been previously suggested to have a protective role in neurodegenerative conditions^34, 35^. Next, we applied our cPP pipeline to analyze the control and PD midbrain dataset published by Smajić et al.^14^ to see if we could find the same disease-associated population in another single-cell study. We found a population of disease-associated ODC exhibiting similar differential expression of disease-associated genes, supporting our result that identifies differentially affected subpopulations of cells in PD (Extended Data Fig. 5c,d).

**Fig. 5.**
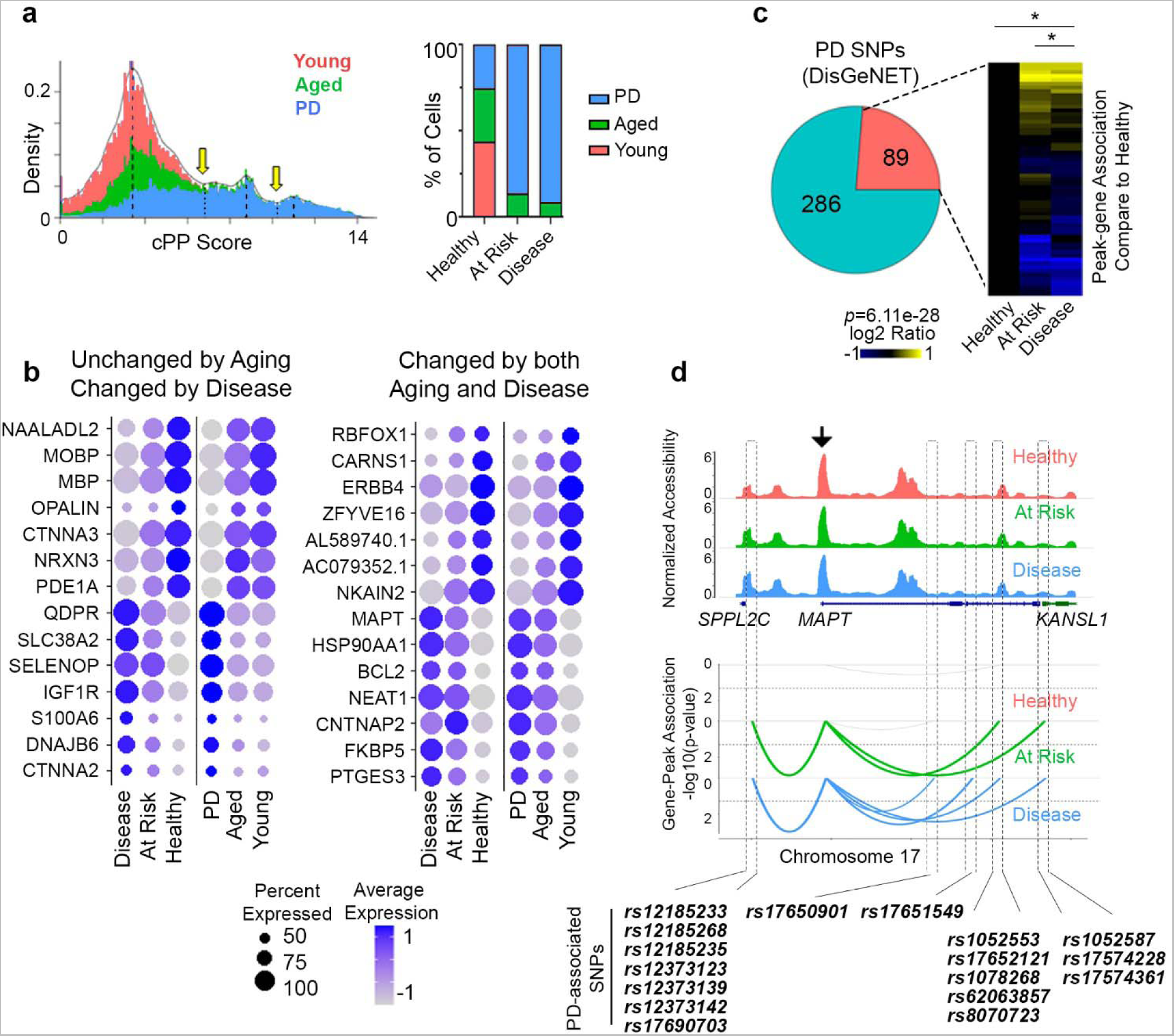
Analysis of disease-associated ODC. **a**, Histogram and inferred multimodal distribution of ODC across cPP. Mode valleys were identified using the multimode package in R. Peaks were identified as healthy, at-risk, and disease-associated ODCs. Bar graph shows percentage of nuclei in each group from the three donor cohorts. **b,** Dot plot of selected genes with differential expression between healthy, at-risk, and disease-associated cells. Left – genes with disease-specific changes with no differential expression between Young and Aged/PD. Right – Genes differentially expressed over aging/PD. **c,** Pie chart representing ratio of PD-associated SNPs within ATAC peaks in our dataset. Pink and cyan color represent the number of PD-associated SNPs inside and outside of our ATAC peaks, respectively. PD-associated SNPs are enriched in our ATAC peaks (LD Score regression, p=6.11e-28). **d,** Peak-gene association plots for peaks with PD-associated SNPs that are significantly correlated with MAPT gene expression in healthy, at-risk, and disease-associated ODC. Black arrow indicates the location of MAPT promoter. SNPs associated with each peak are shown below.

We performed peak-gene analysis of healthy, at-risk, and disease-associated ODC and identified differential peak-gene associations between these groups. Interestingly, 89 previously reported PD-associated SNPs^36^ were located in ATAC peaks that were differentially associated with gene expression in at-risk and disease-associated ODC compared to healthy ones (Fig. 5c). The ATAC peak containing *rs11248060* and *rs11724804* was associated with expression of FGFRL1 and PDE6B in healthy cells, while it was associated with an antisense transcript of undescribed function (AC139887.4) in disease-associated ODC. The peak containing *rs557074* showed no significant peak-gene correlation in healthy ODC but was associated with gene expressions in disease-associated ODC (Extended Data Fig. 5e,f). Of particular interest in this second category, we found five separate peaks near the MAPT locus that contain 17 individual PD-associated SNPs. None of these peaks were associated with MAPT expression in healthy cells, but all five peaks were associated with expression in disease-associated nuclei (Fig. 5d).

To confirm our bioinformatics findings, we performed RNA-FISH on FFPE human substantia nigra sections. As expected, we observed incremental reductions in expression of MBP and CARNS1 over aging and PD (Fig. 6a,b). RBFOX1 expression was decreased in aged controls compared to young subjects (Fig. 6c,d). We also confirmed significant decrease in PDE1A expression (Fig. 6e,f) while SELENOP and QDPR were significantly elevated in PD compared to age-matched controls (Fig. 6g-j). Of note, we saw wide variation in the levels of these genes, even in cells from the same donor, supporting the idea that subsets of cells are differentially affected during the disease process.

**Fig. 6.**
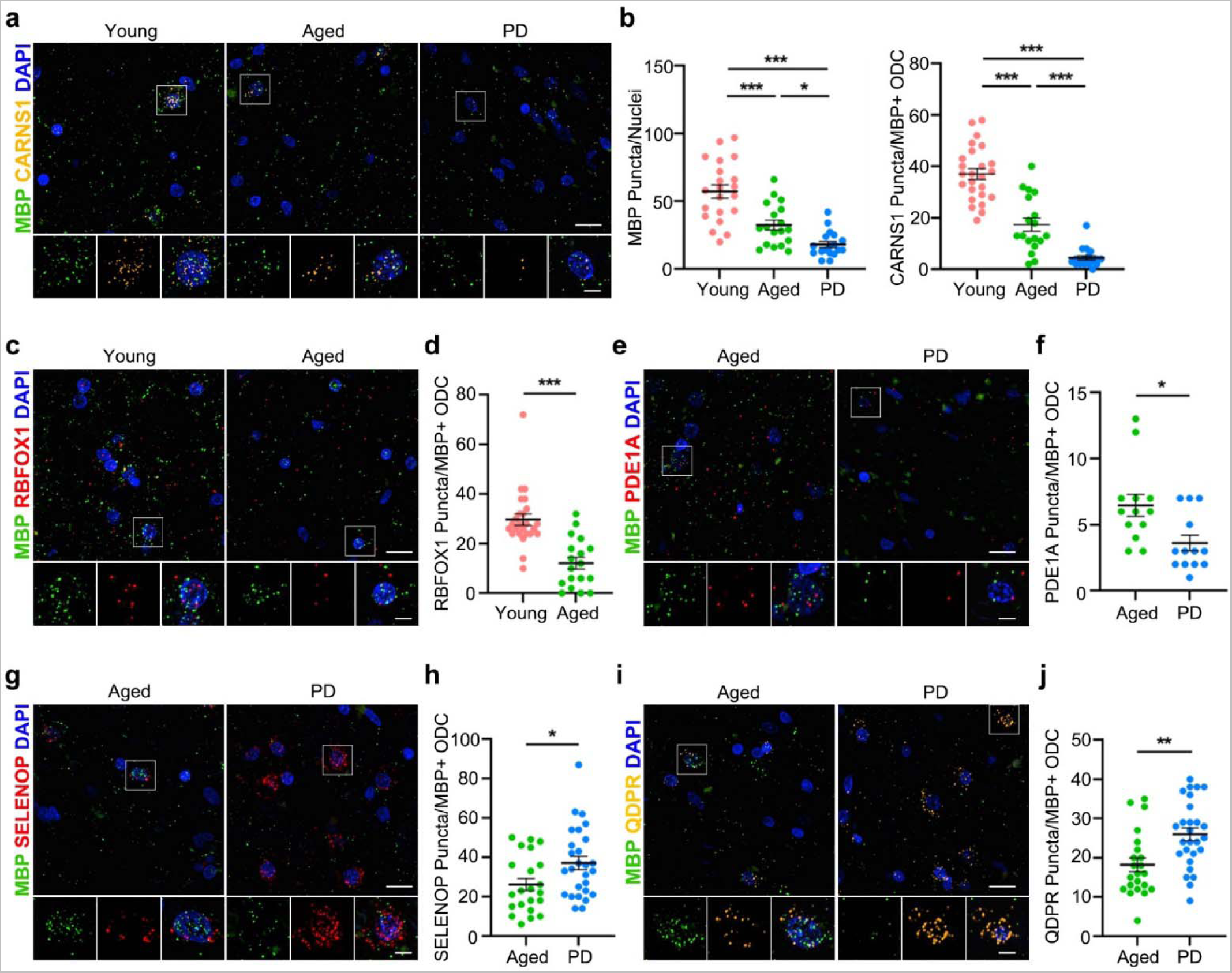
RNA-FISH of human midbrain samples. Confocal imaging (40x) of RNA-FISH for FFPE human midbrain sections for indicated targets; Scale bar = 15µm. Inset (100x) Scale bar = 5µm. **a**, Representative fluorescence images of CARNS1 and MBP showing reduced expression over aging/PD. **b**, Quantification of MBP and CARNS1 puncta within 100x100 pixel square around MBP+ ODC nuclei (One-way ANOVA with Tukey’s post-hoc test. MBP, p-value for Y/A < 0.0001, p-value for A/P = 0.0387; CARNS1, p-value for Y/A < 0.0001, p-value for A/P = 0.0003). **c,** Representative fluorescence images of RBFOX1 showing reduction in aged control compared to young samples. **d,** Quantification of RBFOX1 puncta in MBP+ nuclei (Student’s t-test, p-value < 0.0001) **e,** Representative fluorescence images of PDE1A showing decrease in PD compared to aged control. **f**, Quantification of PDE1A puncta in MBP+ nuclei (Student’s t-test, p-value = 0.0106). **g,i,** Representative fluorescence images ofSELENOP (**g**) and QDPR (**i**) showing increases in PD compared to aged control. **h,j,** Quantification of SELENOP (**h**) and QDPR (**j**) puncta in MBP+ nuclei (Student’s t-test. SELENOP, p-value = 0.0231; QDPR, p-value = 0.0021). For each target, we selected 3-5 random fields from 3 unique donors from each cohort. Independent student t-test, p-value = (*0.05<**0.01<***0.001). Error bars represent SEM.

## Discussion

In this study, we assessed paired snRNA-seq and snATAC-seq patterns in the SN region of the midbrain of young, aged controls and PD patients to elucidate how aging processes may predispose an individual to PD. By adding data from young control donors to the current single-cell studies of the brain, our study may significantly extend our understanding of the aging process and neurodegeneration at single-cell resolution. Leveraging paired analysis of RNA expression and chromatin accessibility from the same cells provides a unique opportunity to identify cell-type specific regulation of genes and their contribution to disease processes. We found there are dramatic and widespread changes in links between expressed genes and discreet chromatin regions during aging and PD. Trajectory analysis revealed all cells change over the course of aging, and there are significant further alterations in ODC and microglia populations in PD, further supporting the recently highlighted roles of these cells in PD development^11–14^. We identified a population of disease-associated ODC and found that even in PD, a large percentage of cells maintain a normal, healthy transcriptional profile. We highlighted genes that were specific for PD but were unchanged over aging and also identified genes that were incrementally affected by aging and PD. These aging-related and disease-associated genes may represent a source of risk that is inherent in the aging process.

Each brain region has a unique cell-type composition. Our data agrees with previous data^11^ indicating that ODC is the major cell type in the midbrain, representing approximately 75% of all cells, which has led to increasing recognition of ODC as a potential major player in PD^11–14^. After ODC, MG, OPC and AS are the next most common cell types; however, we failed to faithfully recover neuronal populations that were reported in previous studies in the midbrain^14^, potentially due to their increased sensitivity to nuclear permeabilization for ATAC-seq preparation.

We adapted the peak-gene association analysis developed by Ma *et al*.^8^ to link distal peaks to genes in *cis* based on co-variation in chromatin accessibility and gene expression, which is a crucial step forward in unraveling the complex cellular heterogeneity of the brain. The recent work by Morabito *et al*.^37^ elegantly combined separate ATAC and RNA expression data to form correlative relationships between two similar nuclei. Peak-gene association studies further advance this technique by analyzing paired data from the same nuclei. The importance of this approach is highlighted by our unexpected findings showing surprisingly little variation in ATAC peaks during aging or PD regardless of changes in gene expression. Despite this, peak-gene analysis revealed widespread alterations in gene expression-peak relationships during aging and PD pathogenesis. For example, we identified the aging-associated lncRNA, NEAT1, is linked to EGR1/2 motif-containing peaks, a downstream NEAT1 effector^38^, even though there were no noticeable changes in chromatin accessibility of regions containing this motif during aging. A more in-depth study of these *cis-*regulatory mechanisms underlying functional alterations in cellular subpopulations is necessary to understand the enormously complex changes that occur during aging and neurodegeneration.

Adapting pseudotime to infer pathogenic changes is a critical step toward expanding our understanding of how aging may alter the midbrain and how those alterations predispose an individual to PD. Our inclusion of young donors with no neurological disease allows us to define a baseline, or ‘healthy’, transcriptional state. Multiomic analysis indicates that ODC and microglia in the midbrain undergo significant population shifts during aging into PD and show an increase in markers previously associated with aging and neurodegenerative disease. We did not observe a strong correlation of midbrain astrocytes or OPC with PD development, although this does not preclude them from having important roles in other brain regions in PD.

Pseudopathogenesis analysis revealed that the majority of ODC in the midbrain maintain their normal functions during aging and PD. However, a small subset of disease-associated ODC with distinct transcriptional signatures appears during the disease process, suggesting that a small number of highly dysregulated cells may drive the major disease process. There were sets of genes selectively changed in PD that were independent of aging such as QDPR and SELENOP, but more importantly, we identified genes that changed over aging that were further altered during PD. As the primary risk factor for PD is aging, these genes may help us understand the difference between so-called ‘healthy’ aging and aging-dependent risk factors for neurological diseases. Of particular note is CARNS1 that encodes carnosine synthase 1. Carnosine (β-alanine-L-histidine) is an endogenous antioxidant and neuromodulator and has been shown to confer protective effects in various neurological conditions including Alzheimer’s disease and PD^34, 35^.We observed that both expression levels and the number of ODC expressing CARNS1 were incrementally decreased over aging and PD. It will be interesting to investigate if the loss of CARNS1 expression in the subpopulation of ODC leads to PD progression. This opens the discussion that oligodendrocytes might play a key role outside of their canonical myelination function to maintain neuronal health, and that loss of this support may tip the balance of the highly vulnerable dopaminergic neurons into a degenerative state.

Taking advantage of our paired multiomic dataset allows us to contextualize the information provided by PD GWAS. We identified that ATAC peaks containing 89 known PD-associated SNPs showed significant alterations in peak-gene associations between disease-associated cells and transcriptionally normal cells. It is imperative to obtain functional inferences of these PD-associated SNPs in differential gene regulation toward PD pathogenesis.

In future studies, a more nuanced gradient of aging samples may be required to understand the continuum of cell states and how populations shift during aging. It will also be interesting to see what findings of similar studies in different brain regions will uncover, as this is essential to building a brain-wide atlas of human aging and neurodegeneration.

## Methods

### Nuclei isolation and sequencing

Human tissue samples were obtained through the NIH Neurobiobank from the Human Brain and Spinal Fluid Resource Center (VA Greater Los Angeles Healthcare System). To capture the SN region of the midbrain precisely, we dissected approximately 3 x 3 x 5 mm sections using postmortem photos provided (Supplementary Fig.1a), yielding tissue blocks ranging from 50-75 mg. We processed the tissue in a dounce homogenizer and then centrifuged it through 1.5 M sucrose to isolate the nuclei. With this method, we are able to reliably isolate > 1x10^6^ intact nuclei from ∼50 mg postmortem midbrain tissue. Centrifugation through an iodixanol gradient as previous described^9^ produced high quality nuclei with low mitochondrial contamination, but the actual yield of nuclei was very low (2.5-5x10^4^ nuclei from ∼50 mg postmortem midbrain tissue), so we did not continue with this method as the SN is a small region and tissue amount is limited. We stained the nuclei with 7AAD and sorted to purify nuclei from mitochondria. After sorting, we permeabilized according to current 10XGenomics protocols (Protocol CG000375 Rev A: 10 mMTris-HCl (pH 7.4), 10 mM NaCl, 3 mM MgCl_2_, 0.1% Tween-20, 0.01% Digitonin, 1% BSA, 1 mM DTT, 1 U/uL RNase Inhibitor) for 2 min on ice). Nuclei were washed once before counting and proceeding with 10XGenomics protocols for transposition, nuclei isolation and barcoding, and library preparation exactly as written (10XGenomics Protocol Chromium Next GEM Single Cell Multiome ATAC + Gene Expression CG000338 Rev A). Libraries were sequenced at GeneWiz using Illumina Novaseq S4 flowcells. Our actual average sequencing depth was 1.8E8 reads/sample for gene expression libraries and 1.75E8 reads/sample for ATAC libraries. After sequencing, three samples (one each of Young, Aged, and PD) showed strong ATAC results but poor RNA read quality indicative of RNA degradation in the original sample. The nuclei from these three donors were excluded from downstream analysis.

### Authors’ note on nuclear isolation

This isolation method was sufficient to obtain high quality nuclei with minimal blebbing and worked well in our initial pilot study. However, after in-depth data analysis, we noted that barcodes for nuclei with neuronal expression patterns consistently showed unusually high RNA read count, suggesting that these nuclei were clumping together and forming multiplets, and thus, these neurons were excluded by our QC filtering (see below). In nuclear isolations this is commonly caused by a partial loss of membrane integrity that allows genomic DNA to leak out and cause nuclei to stick together. We feel it is likely the digitonin permeabilization step may have been too harsh for these samples, and that the larger neuronal nuclei were more sensitive to digitonin. Previous publications have also noted the effect of differential responses in cell types to different isolation protocols^6^. In the months since we performed the initial isolation, multiple alternative protocols for isolation of frozen post-mortem tissue have been posted and shared online in unofficial forums that omit digitonin for this reason. In future studies, we will also adjust our isolation protocols accordingly.

### Quality control of single-nuclei RNA+ATAC multiome

Both single-nuclei RNA and ATAC-seq reads were spontaneously mapped to GRCh38 human reference transcriptome and genome, respectively, by Cell Ranger ARC (v1.0.1) with ”--mempercore=8 --localcores=12” (https://support.10xgenomics.com/single-cell-multiome-atac-gex/software/downloads/latest). Quality control (QC) of single-nuclei transcriptome and chromatin profiles was conducted by Seurat (v4.0.0) and Signac (v1.1.1) in R package (v4.0.2), respectively^16, 39^. Briefly, we removed cells with 1) less than 100 or more than 7,000 detected genes, 2) less than 500 or more than 20,000 reads and 3) more than 5% mitochondria-derived reads (damaged/dead cells or doublet) in the RNA datasets. We also evaluated the doublet frequency by counting cells expressing both *STMN2* and *AQP4* (Fig S1d), which are usually exclusively expressed in cortical neuron and astrocyte, respectively^40, 41^. If the ratio of *STMN2^+^AQP4^+^* cells to total number of *STMN2^+^AQP4^-^*, *STMN2^-^AQP4^+^*and *STMN2^+^AQP4^+^* cells was more than 3%, the libraries were excluded from subsequent analyses. For QC of ATAC datasets, we calculated four measurements: 1) ratio of mononucleosomal fragments to nucleosome-free fragments, 2) enrichment score in transcription start site (TSS), 3) total number of ATAC-seq reads per cell and 4) ratio of reads mapped to “blacklist regions” that are genomic regions inducing aberrant mapping and artificial signals^42^. We filtered out cells with 1) more than 4 mononucleosome:nucleosome-free ratio, 2) less than 2 TSS enrichment score, 3) less than 1000 and more than 60,000 ATAC reads, 4) more than 2% of reads in the blacklist regions and 5) less than 1,000 or more than 25,000 ATAC peaks. Our initial isolation yielded 82735 nuclei. After QC we retained 69,289 high-quality nuclei that met the above criteria of RNA and ATAC for subsequent analyses (Fig. S1d-g).

### Integration of single-cell gene expression profiles

Gene expression profiles were harmonized using Seurat (v4.0.0)^3^ as we have done previously^17, 18, 40, 41, 43^. In each snRNA-seq library, raw UMI count was normalized to total UMI count. Highly variable genes were then identified by variance stabilizing transformation with 0.3 loess span and automatic setting of clip.max value. The top 2,000 variable genes were used to identify cell pairs anchoring different snRNA-seq libraries using 20 dimensions of canonical correlation analysis (CCA). After scaling gene expression values across all integrated cells, we performed dimensional reduction using principal component analysis (PCA). For the visualization, we further projected single cells into two-dimensional Uniform Manifold Approximation and Projection (UMAP) space from 1st and 20th PCs. Graph-based clustering was then implemented with shared nearest neighbor method from 1^st^ and 20^th^ PCs and 0.8 resolution value. Differentially-expressed genes (DEGs) in each cluster were identified with more than 1.25-fold change and p<0.05 by two-sided unpaired T test. Note:

Due to inherent imbalance in sex of donor samples, differential gene analysis between donor cohorts includes sex-specific genes such as XIST and UTY, which are omitted from downstream analysis. Gene Ontology (GO) analysis was performed to DEGs by GOstats Bioconductor package (v2.56.0)^44^. False discovery rate was adjusted by *p.adjust* function in R with “method=”BH”” option.

### Cluster annotation

Cell types were assigned in each cluster “island” with uniquely-expressed genes^18, 43^. First, we assigned neuronal clusters with expression of *STMN2* and *TBR1.* Oligodendrocyte and astrocyte clusters were classified by myelination markers (*MBP* and *MOG*) and astrocyte-specific proteins (*GFAP* and *AQP4*) expression. *OLIG1^+^OLIG2^+^*clusters without any neuronal and astrocyte markers were defined as oligodendrocyte precursor cells (OPCs). Endothelial cells were annotated by substantial expression of *FLT1*, *VWF* and *PDGFRB*. We annotated clusters with *GPR34*, *TREM2* and *C1QC* without any OPC and astrocyte markers. Since T-cell infiltration into substantia nigra is a hallmark of PD pathology, we also defined T-cell clusters with their specific marker expression (*CD3E* and *CD8A*).

### Integration of single-cell chromatin profiles

Integrative analysis of single-cell chromatin profiles was performed by Seurat (v4.0.0), Signac (v1.1.1), GenomicRanges (v1.42.0) and Harmony (v1.0) R packages^39, 45–47^. First, ATAC peaks from all ATAC libraries were merged by *reduce* function in GenomicRanges. Huge (>10,000bp) and tiny (<20bp) combined peaks were removed from subsequent analyses. ATAC reads were recounted in the combined peaks by using the *FeatureMatrix* function in Signac. After merging the peak x cell matrices from all ATAC libraries, the ATAC read counts were normalized by term frequency inverse document frequency. Latent sematic indexing (LSI) was then computed from the merged count matrix by the singular value decomposition method. To minimize the batch and technical difference across libraries, we ran the Harmony algorithm using LSI embeddings with “project.dim=F” option. For data visualization, all cells were embedded into two-dimensional UMAP space from 2nd and 30th Harmony-corrected LSI. The 1^st^ index of LSI may represent technical variations, such as sequencing depth and was not used for UMAP projection.

### Analysis of differential gene expression and chromatin accessible regions

Gene expression and chromatin accessibility profiles were compared between PD and healthy aged groups, between healthy aged and young groups or across cell types. The differentially-expressed genes and differential open chromatin regions were identified in each cluster by 1.25-fold change and p<0.05 unpaired T test. Cellular events and functions related to the differentially-expressed genes and differential open chromatin regions were analyzed by the enrichment of Gene Ontology using GOstats (v2.56.0) as described above^44^. Genomic distribution of all ATAC peaks and differential open chromatin regions was identified by *annnotatePeak.pl* script in HOMER suite (v4.11.1) with default parameter^48^. Analysis of previous association of differentially expressed genes was done manually^6, 49–72^.

### Analysis of relationship between gene expression and ATAC peaks

To investigate the relationship between regulatory sites and differential gene expression, Spearman correlation was calculated using the normalized gene expression values and ATAC read counts in each gene-peak pair^8^. We chose all ATAC peaks within ± 500kbp of TSSs of genes for the correlation analysis. In each peak, we generated 100 permutated background peaks with the same accessibility. Spearman correlation was also calculated to the background peaks to estimate the background correlation distribution. We assumed that the background correlation follows Gaussian distribution. Thus, we determined p-value of the observed Spearman correlation using *pnorm* function in R with mean and standard deviation of the background correlation distribution. We defined significant gene-peak association with p<0.05.

### Subgrouping and pseudopathogenesis analysis in each cell type

In each cell type, individual cells were re-scaled in both RNA and ATA profiles by SCTransform (v0.3.2). Dimensionality reduction was then implemented by PCA and UMAP embedding by Seurat as described above. Subsequently, we performed cell trajectory analysis using Monocle3 (v0.2.3.0) to estimate pathological stages of individual cells^32^. Briefly, Seurat object was converted into Monocle3 “cell_data_set” format by SeuratWrapper (v0.3.0). Clustering was performed by Leiden community detection. Principal graph of cell trajectory was constructed using *learn_graph* function with default parameter and used for pseudotime calculation by choosing cells in young group-specific clusters as root cells. In each cell *i*, two pathological measurements, RNA- and ATAC-derived pseudopathogenesis (*PP_RNA,i_*and *PP_ATAC,i_*), were combined by dividing their variances of all cells *(σ^2^* and *σ^2^*) as follows:

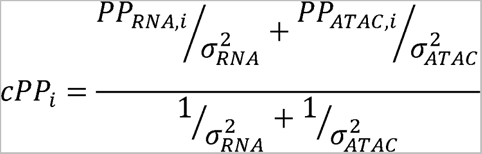

Here, we called the combined values with corrected errors PseudoPathogenesis (c*PP_i_*). To identify differential genes and peaks along pseudopathogenesis, we calculated Spearman correlation with the scaled gene expression and peak intensity values. We chose genes and peaks with > 0.1 or < -0.1 for subsequent analyses. Peaks associated with at least one pseudopathogenesis-correlated genes were used for subsequent motif enrichment analysis.

### Functional subtype gene modules

To identify potential subsets of nuclei with functional differences, we used literature-derived gene sets that have been previously reported^21, 22, 24–26^. For each set of genes, we used the AddModuleScore function of Seurat to establish a combined expression score for each nucleus for all the genes in the list, and added it as metadata. The gene sets for each oligodendrocyte module are as follows: newly formed ODC (*TCF7L2, CASR, CEMIP2, ITPR2*); myelin forming ODC (*MAL, MOG, PLP1, OPALIN, SERINC5, CTPS1*); mature ODC (*KLK6, APOD, SLC5A11, PDE1A*); Synaptic Support ODC (*NFASC, NRXN3, CNTNAP2, ANK3*); ODC-Neuron Adhesion Markers (*HAPLN2, STMN1, MAP1B, SEMA5A, EPHB2, S100B, PRKCA*). After cPP analysis, two additional modules were identified based on cPP-relevant gene expression and GO analysis: Protein Folding (*HSP90AA1, HSPA1A, HSPA1B, CRYAB, FKBP5*) and Chaperone Mediated Autophagy (*LAMP2, ST13, DNAJB1, STIP1, HSPA8, BAG3*). The gene sets for microglia modules are as follows: homeostatic MG (*HEXB, CST3, CX3CR1, CTSD, CSF1R, CTSS, SPARC, TMSB4X, P2RY12, C1QA, C1QB*); stage 1 TREM2 independent DAM MG (*TYROBP, CTSB, APOE, B2M, FTH1*); stage 2 TREM2 dependent DAM MG (*TREM2, AXL, CST7, CTSL, LPL, CD9, CSF1, ITGAX, CLEC7A, LILRB4, TIMP2*); and Aging MG (*IL15, CLEC2B, DOCK5*). The gene sets for Astrocyte modules are as follows: Disease-Associated AS (*GFAP, CSTB, VIM, OSMR, GSN, GGTA1P*); A1 Reactive AS (*SERPING1, GBP2, FKBP5, PSMB8, SRGN, AMIGO2*); A2 Reactive AS (*CLCF1, TGM1, PTX3, S100A10, SPHK1, CD109, EMP1, SLC10A6, TM4SF1, CD14*), and GFAP-low AS (*LUZP2, SLC7A10, MFGE8*).

### Cell Classification by cPP

To infer multimodality of cPP histogram, we performed Kernel density estimation by multimode R package^33^. Briefly, we applied locmodes function to cPP scores of ODC with “mod0=2” and estimated the location of anti-modes (valleys) of multimodal distribution. We used anti-modes between first and second modes and between second and third modes as thresholds of “At Risk” and “Disease” group, respectively. Comparison of previously published single-cell RNA-seq data^14^ was done performed using the same analysis pipeline previously described for oligodendrocytes. Briefly, the ODC subset was processed using SCTransform with the previously defined parameters. To identify if there is also a clear trajectory from healthy to disease states, we used the top 10 differentially expressed genes between healthy and disease to establish a gene expression module. Using the clusters with the highest expression of the healthy and disease gene modules as the root cells, we constructed a principal graph of cell trajectory using *learn_graph* function with default parameters. We noted the appearance of a single large mode peak at the highest pseudotime value and denoted these nuclei as Disease for the purposes of comparative analysis. We also applied the same analysis technique to other glial cell types from our samples, but the smaller cell populations tend to generate multiple small histogram peaks and we elected not to include this analysis to prevent erroneous conclusions from being drawn through over interpretation or ‘force-fitting’ of data.

### Motif enrichment analysis

Transcription factor binding motifs in ATAC peaks were identified using the HOMER (v4.11) suite. Briefly, we create BED format files for specific sets of ATAC peaks and ran *findMotifsGenome.pl* script with “-size given” option to evaluate statistical significance of the motif enrichment. -log10(FDR) value was used as a score of motif enrichment in the set of ATAC peaks.

### SNP-containing peaks

Curated PD-related SNPs were obtained from DisGeNET database^36^. PD-related SNP loci within ATAC peaks were detected by BEDtools^73^. To evaluate statistical significance, we calculated expected frequency of PD-related SNPs by randomly subsampling genomic regions, whose number and size are the same with ATAC peaks. We repeated the subsampling at 10 times and estimated Gaussian probability distribution. Then p-value of the observed count of PD-related SNPs within ATAC peaks were calculated from the estimated probability.

### Single-Molecule RNA-FISH

Multiplex RNA-FISH in 5µm-FFPE human substantia nigra brain tissue sections was performed using RNAscope Multiplex Fluorescent V2 Assay kit (Advanced Cell Diagnostics) according to manufacturer’s instructions. Probes were designed and manufactured by Advanced Cell Diagnostics; Hs-CARNS1-C1(1171001), Hs-PDE1A-C1(446121), Hs-SELENOP-C1(512831), Hs-QDPR-C2(560001), and Hs-MBP-C3(411051).

Sections were imaged using Andor Dragonfly 2000 confocal with Nikon 40X and 100X objectives. Quantification of RNAScope signal was performed using the Imaris (v9.9) imaging suite ‘Dots’ functionality (see Supplementary Fig 11). Cells with nuclear MBP signal were considered to be oligodendrocytes. We counted all positive RNAScope puncta over 0.75um in diameter within a 100 pixel (18µm x 18µm) square centered around the oligodendrocyte nuclei, for the full depth of the tissue slice (5µm). We used 3 donors from each cohort and selected 3-5 random fields (based on tissue size and condition) from each tissue slide for analysis.

## Data Availability

Processed data for the samples presented in this study is available in the Gene Expression Omnibus (GEO) database under accession number GSE193688. Raw data will be made available through the dbGap portal, and specific access information will be added once the deposit request has been finalized.

https://www.ncbi.nlm.nih.gov/geo/query/acc.cgi?acc=GSE193688

## Data Availability

https://www.ncbi.nlm.nih.gov/geo/query/acc.cgi?acc=GSE193688

## Acknowledgement

Authors gratefully acknowledge NIH Neurobiobank for providing all postmortem brain samples. Authors also thank Mr. Andrew Knott for language editing.

## Author contributions

L.A. and Y.S.K designed the study. Nuclear isolation and library preparation were performed by L.A. and M.S. Computational analysis was performed by Y.T. and L.A. L.A., Y.S.K., and Y.T. wrote the manuscript; all authors contributed to review and revision of the manuscript.

## Human Samples

Human midbrain samples were obtained through NIH NeuroBioBank requests (The Human Brain and Spinal Fluid Resource Center, University of Maryland Brain and Tissue Bank, Harvard Brain Tissue Resource Center, University of Miami Brain Endowment Brank). This project utilized de-identified post-mortem brain samples, so is not considered to meet federal definitions for IRB jurisdiction, and falls outside the purview of the Rutgers IRB committee.

---

**Extended Data Fig. 1.**
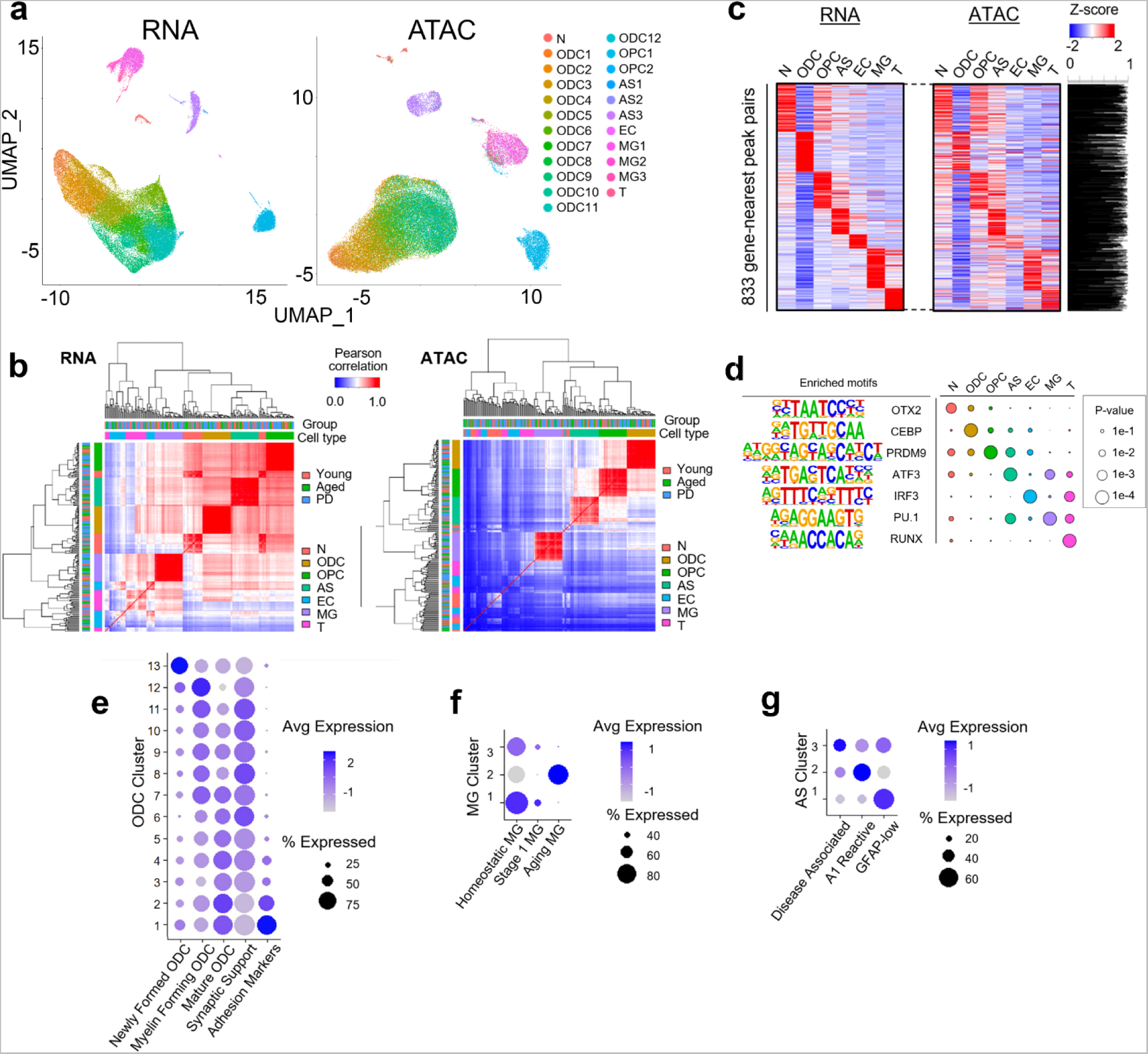
Multiomic analysis of human midbrain. **a**, UMAP visualization of single nuclei by RNA (left) and ATAC (right) profiles. Nuclei are colored by 23 joint clusters. **b,** Heatmap showing Spearman correlation of average RNA expression (left) and ATAC peaks profiles (right) by cell types for each individual. Top and second color bar represents groups of donors and cell types, respectively. **c,** Heatmap showing RNA expression and ATAC peak intensity of cell type-specific genes and their nearest peaks, respectively. ATAC peak intensity is calculated in the nearest peaks from TSS. Pearson correlation across cell types in each gene-peak pair is shown in the right panel (Cor=0.797±0.226). **d,** Enrichment of motifs in each annotated cell type. **e-g,** Dot plot displaying enrichment of gene expression modules for functionally distinct subpopulations for ODC **(e)**, MG **(f)** and AS **(g)** clusters in the human midbrain.

**Extended Data Fig. 2.**
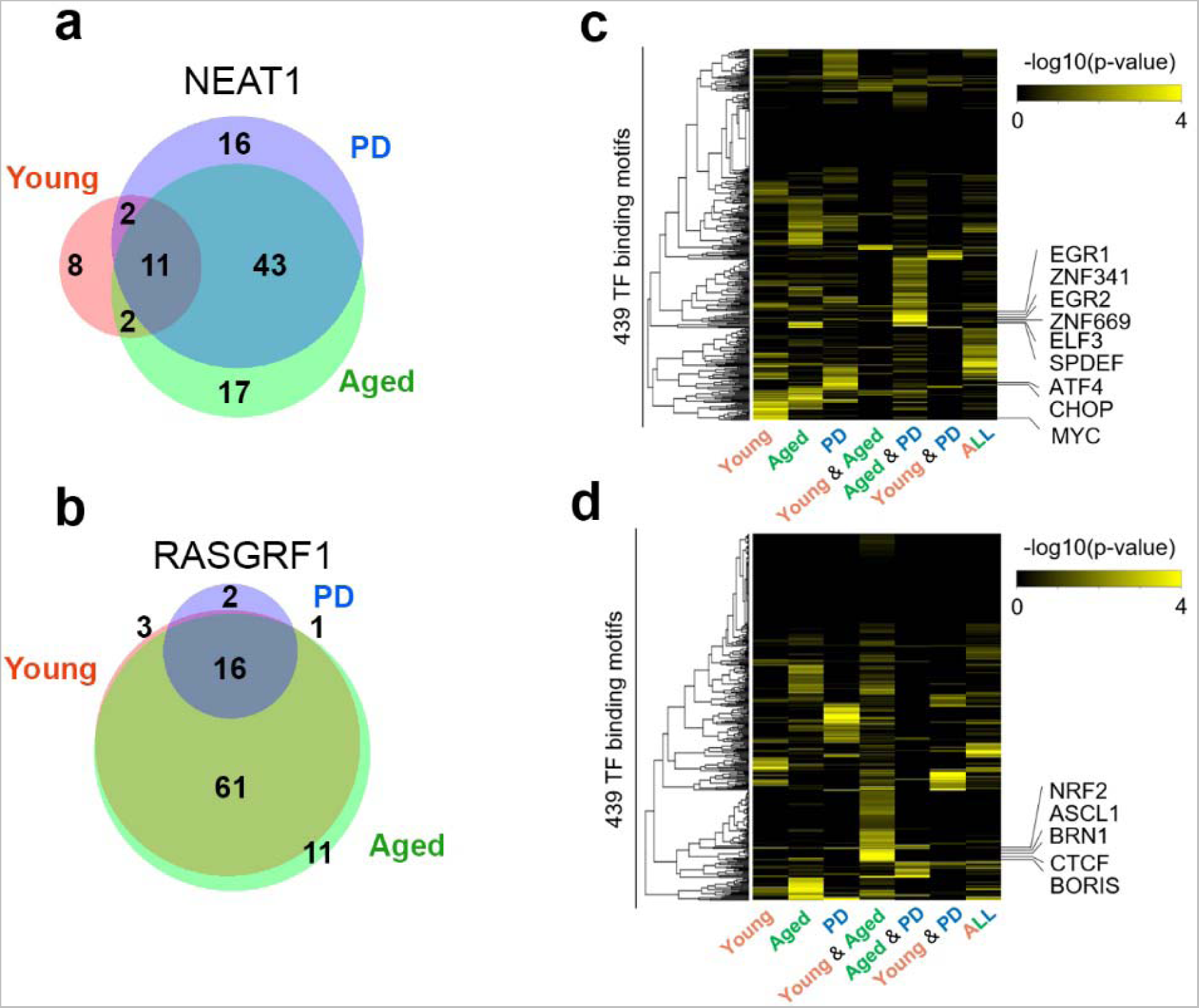
Changes in gene-peak connections in young, aged, and PD for NEAT1 and RASGRF1. **a,b**, Venn diagram of the number of associated peaks with NEAT1 **(a)** and RASGRF1 **(b)** for young, aged and PD midbrain. **c,d,** Heatmap showing enrichment of TF binding motifs in associated peaks with NEAT1 **(c)** and RASGRF1 **(d)**.

**Extended Data Fig. 3.**
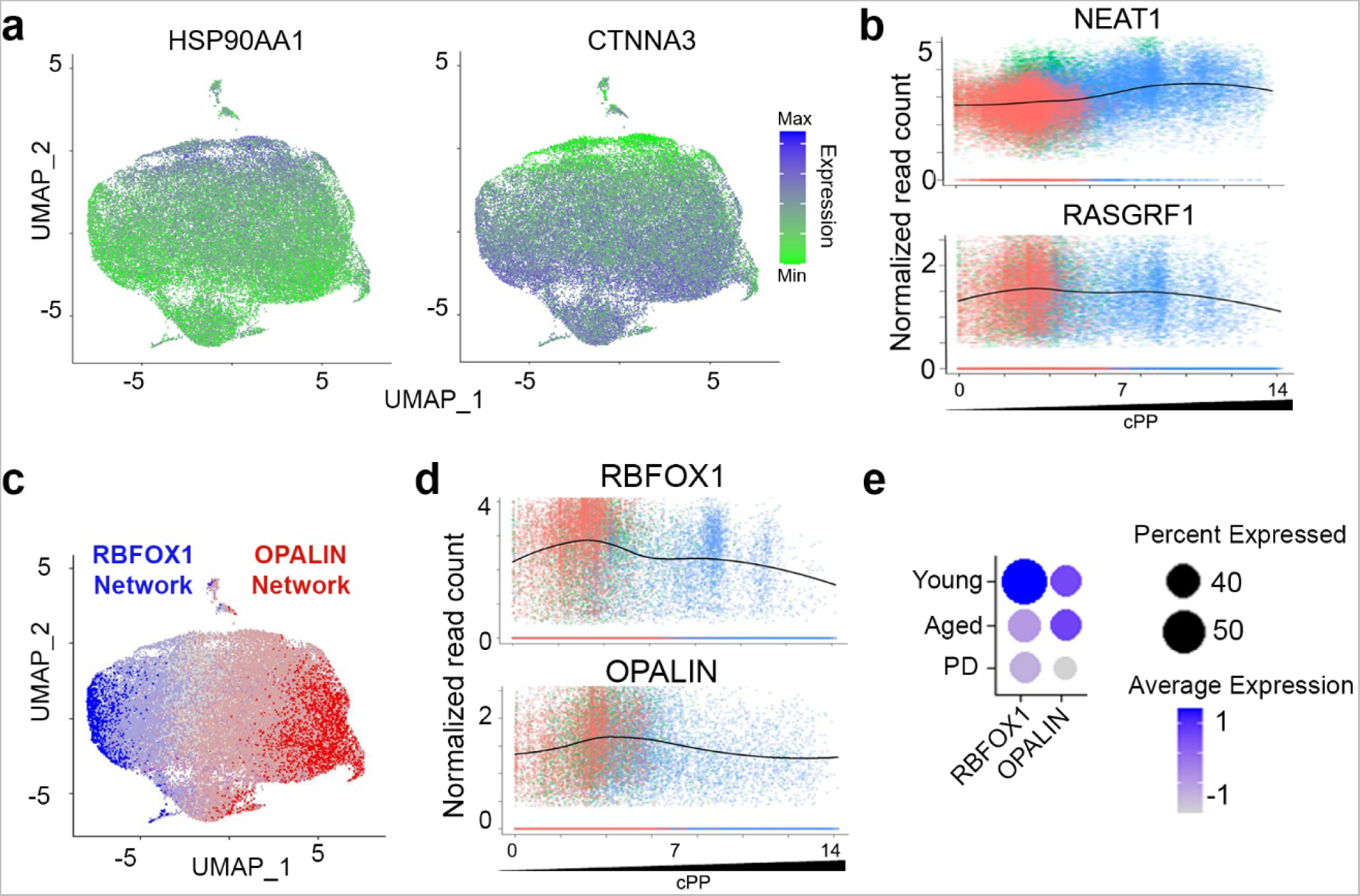
cPP analysis of ODC. **a**, Example genes that have correlated increase (HSP90AA1) and decrease (CTNNA3) of expression with cPP trajectory. **b,** Plot of expression of NEAT1 and RASGRF1 across cPP trajectory. NEAT1 expression is correlated with increasing cPP score; RASGRF1 expression is inversely correlated with cPP score (Cor > 0.1 or < -0.1). **c,** UMAP of RBFOX1 and OPALIN and their coexpressed genes are mutually exclusive in ODC. **d**, Expression plot of RBFOX1 and OPALIN across cPP trajectory. **e**, Dot plot of gene expression for RBFOX1 and OPALIN in each donor cohort.

**Extended Data Fig. 4.**
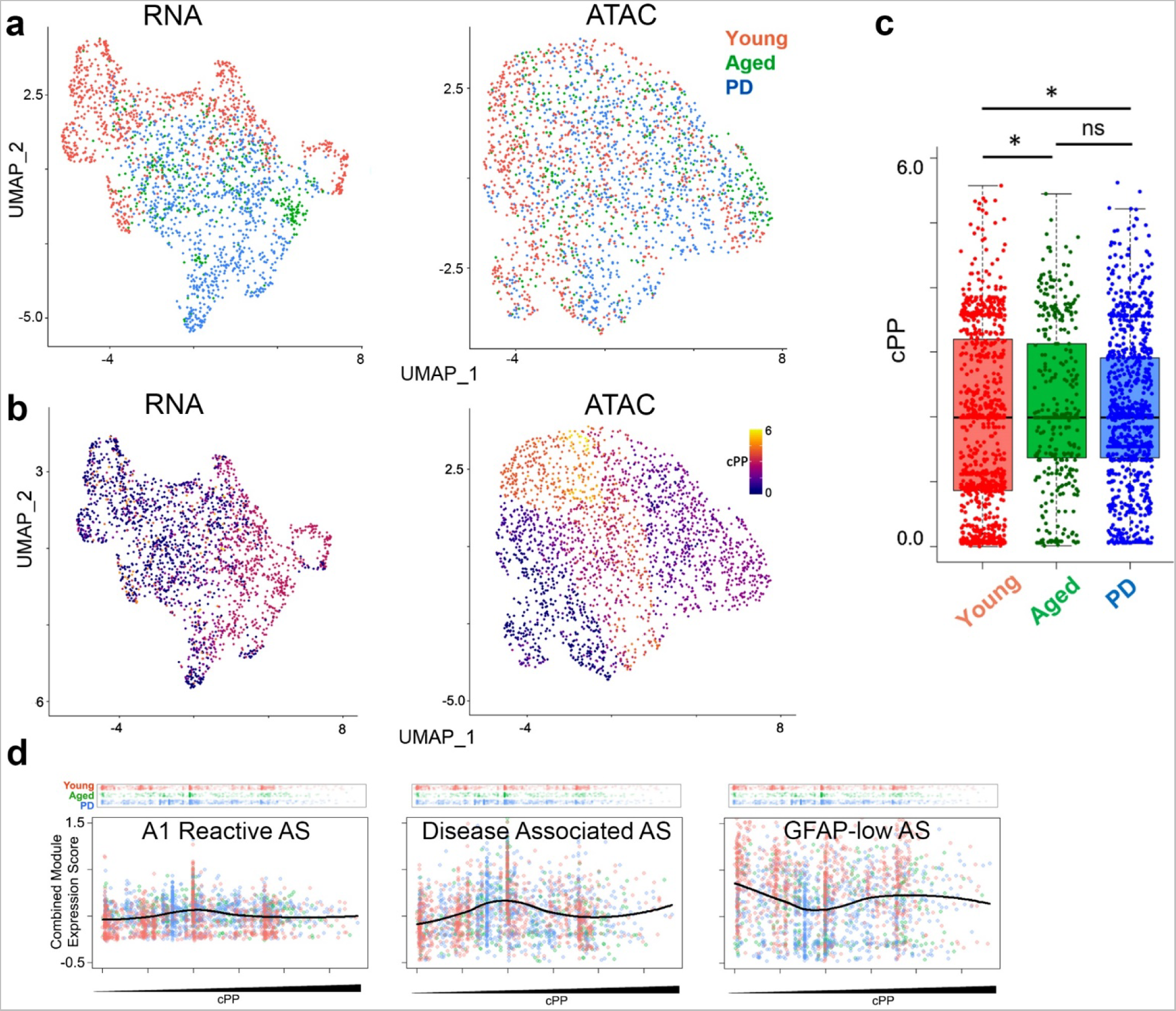
Establishment of pseudopathogenesis trajectory in AS. **a**, UMAP plot of AS nuclei colored by young, aged and PD donor. **b,** UMAP plot of AS nuclei colored by cPP. **c,** cPP scores of individual AS nuclei from young, aged and PD midbrain are significantly changed over aging but not disease state. (One-way ANOVA, p-value for Y/A = 0.002, p-value for A/P = 0.24). **d,** Gene expression modules across AS cPP trajectory. Top panel shows individual nuclei AS along cPP scores and donor group. X-axis shows cPP score. Y-axis is combined expression level for all genes in the expression module.

**Extended Data Fig. 5.**
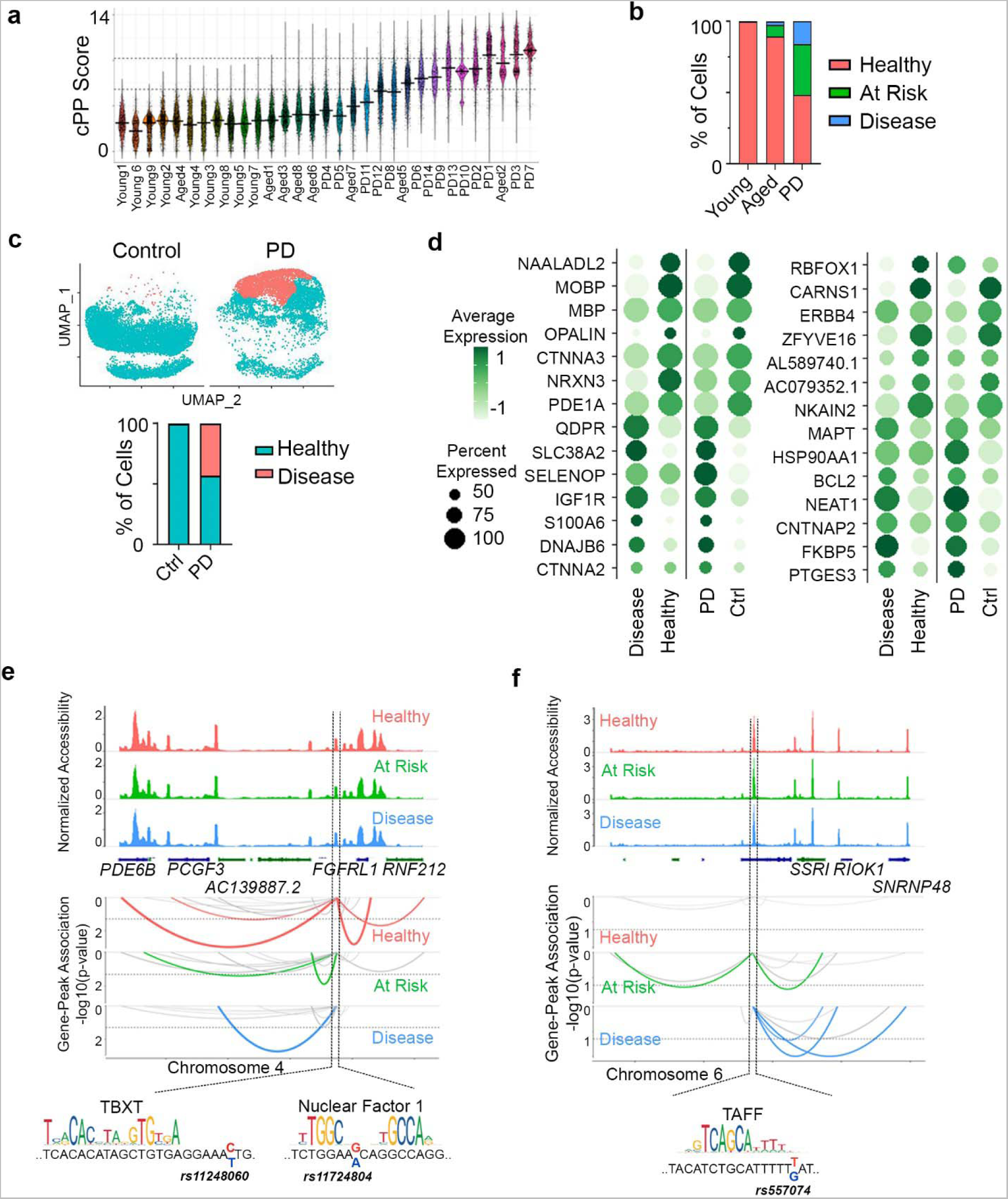
Aging- and disease-specific analysis. **a**, Violin plot of cPP score of every individual ODC nuclei by donor. **b,** Bar graph showing percentage of nuclei in donor cohort from healthy, at-risk, and disease groups. **c,** UMAP showing healthy and disease subsets from publically available snRNA-seq data (Smajić et al, 2022) from the human PD and aged control midbrain after pseudopathogenesis analysis. **d,** Dot plots of genes that from the same dataset (Smajić et al, 2022) showing similar expression patterns between healthy and disease as our multiomic dataset. **e,f**, Representative peak-gene connection plots for peaks containing PD-associated SNPs that have decreased (**e**) or increased (**f**) gene connections in disease-associated ODC compared to healthy ones. Motif information was obtained from JASPAR Transcription Factors track in the UCSC genome browser. SNPs associated with each peak are shown below.

