## Supplementary Figures for "Single-nuclei paired multiomic analysis of young, aged, and Parkinson’s disease human midbrain reveals age- and disease-associated glial changes and their contribution to Parkinson’s disease"

**Supplementary Figures 1-10 for:**

**Single-nuclei paired multiomic analysis of young, aged, and Parkinson’s disease human midbrain reveals age-associated glial changes and their contribution to Parkinson’s disease**

Levi Adams^1†^, Min Kyung Song^1†^, Yoshiaki Tanaka^2✉^, Yoon-seong Kim^1✉^

1 – RWJMS Institute for Neurological Therapeutics, Rutgers-Robert Wood Johnson Medical School, Piscataway NJ, USA

2 - Maisonneuve-Rosemont Hospital Research Center (CRHMR), Department of Medicine, University of Montreal, QC, Canada

† These two authors contributed equally.

✉ Corresponding authors

Yoon-Seong Kim, M.D., Ph.D., RWJMS-Institute for Neurological Therapeutics at Rutgers

683 Hoes Lane West, Piscataway, NJ 08854

Yoshiaki Tanaka, Ph.D., Maisonneuve-Rosemont Hospital Research Center (CRHMR), Department of Medicine, University of Montreal, QC, Canada

5415 boulevard de I'Assomption, Montreal, Quebec, H1T 2M4, Canada

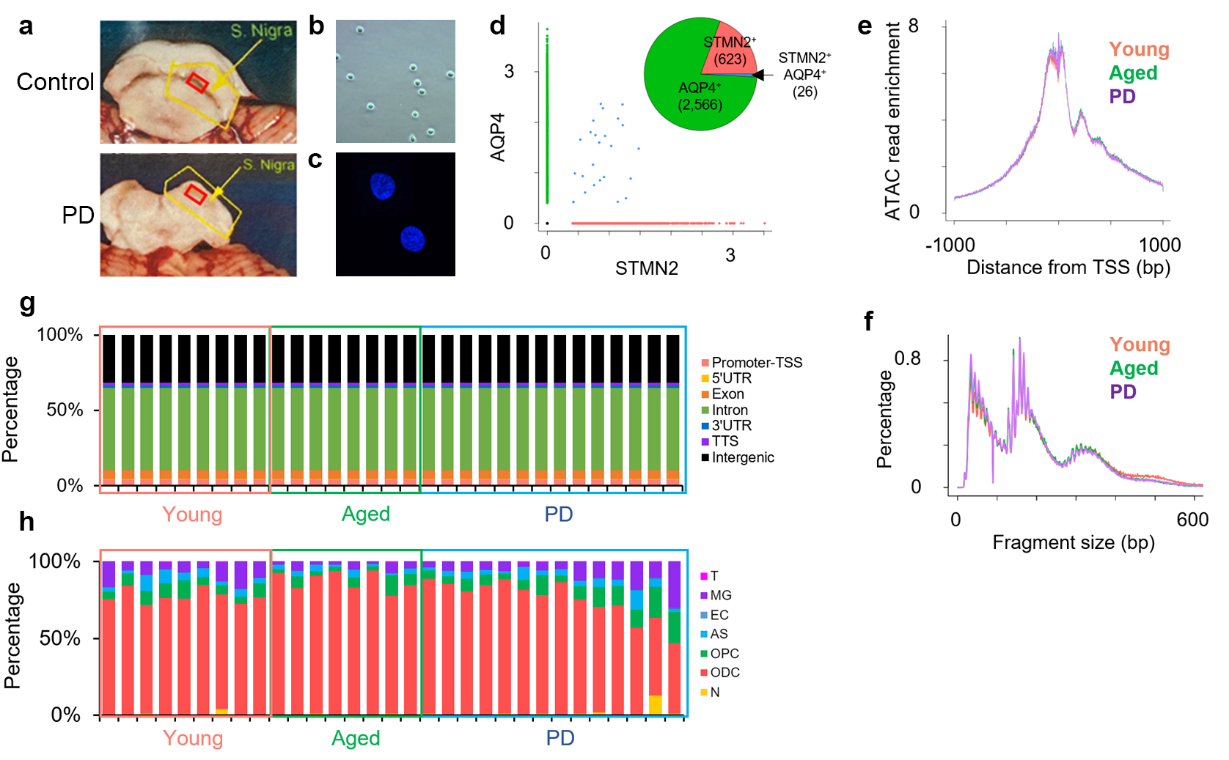

**Supplementary Fig. 1 | Sample preparation and quality control. a,** Photograph of post-mortem midbrain containing the substantia nigra with sampled area highlighted in red. **b,c,** Microscope images of actual nuclei obtained after isolation and permeabilization with phase-contrast at 20x **(b)** and with Hoechst staining at 100x **(c)**. **d,** Comparison of mutually exclusive gene expression shows < 0.1% overlap. **e,** Enrichment of ATAC reads around transcription start sites (TSS). Y-axis represents normalized ATAC read count. **f,** Histogram of fragment size of ATAC reads. **g,** Percentage of ATAC peak distribution. Each bar represents one donor. **h,** Percentage of high-quality nuclei for each individual cell type. Each bar represents one donor.

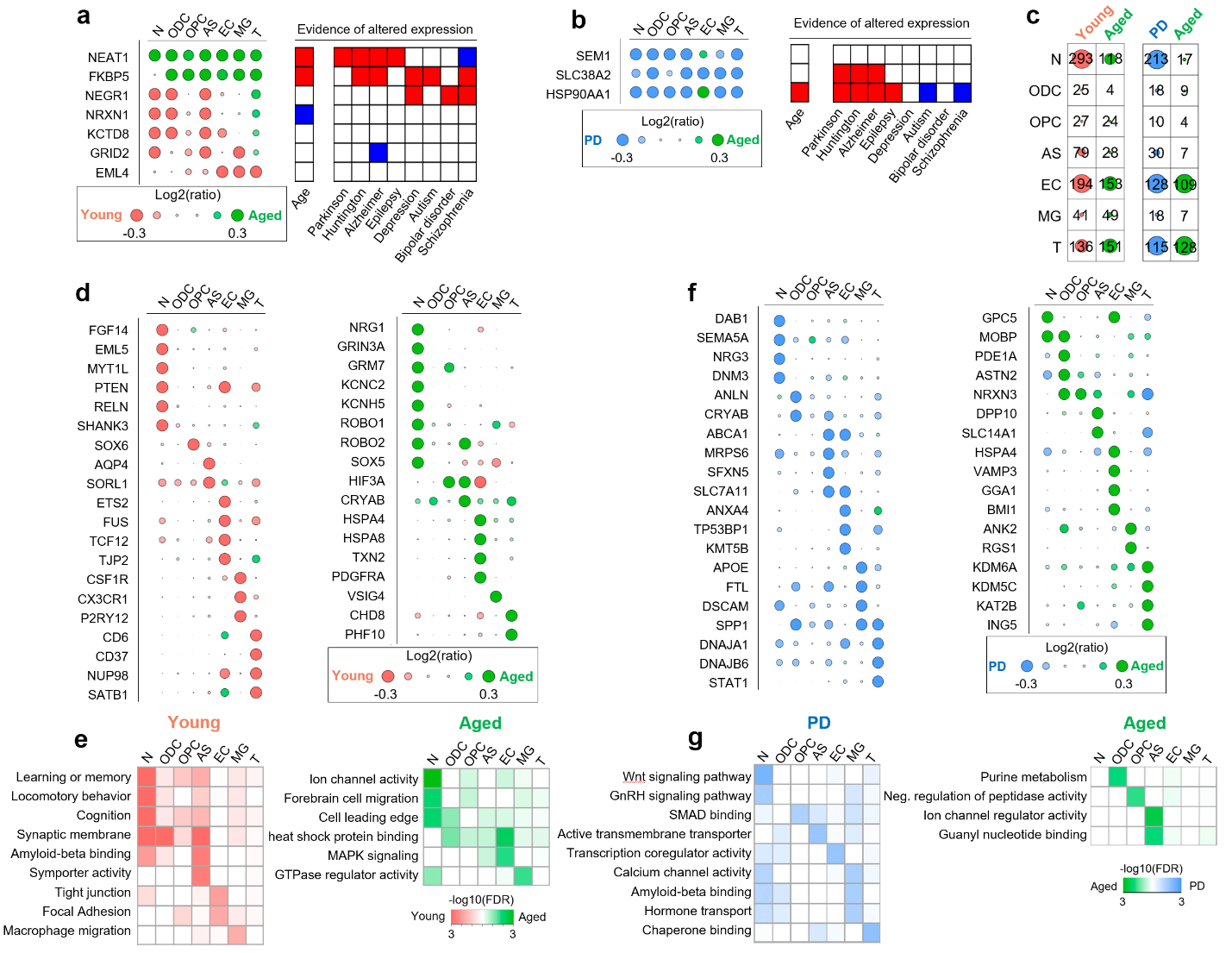

**Supplementary Fig. 2** **| Differential gene expression between young, aged and PD midbrain.** **a,b,** Circle plot showing differentially-expressed genes between young and aged **(a)** and between PD and aged **(b)** in more than three cell types. Literature-based evidence^1-27^{Zhao, 2019 #549}{Zhao, 2019 #549;Ximerakis, 2019 #596;Woo, 2017 #574;Uryu, 2006 #572;Sunwoo, 2017 #543;Sinclair, 2013 #557;Sha, 2017 #575;Patel, 2016 #556;Patel, 2019 #565;Mariani, 2016 #562;Maccarrone, 2013 #559;Liu, 2018 #542;Lee, 2011 #571;Labadorf, 2015 #564;Kim, 2001 #576;Katsel, 2019 #552;Karis, 2018 #560;Evers, 2002 #577;Chang, 2014 #558;Blair, 2013 #553;Binder, 2004 #555;Barry, 2015 #541;Barry, 2017 #551;Baldo, 2012 #573;Al-Dalahmah, 2020 #554}{Zhao, 2019 #549} of altered expression by aging and neurological diseases are shown in the right panel. Red and blue color represent up and down regulation in the patient/animal model brain, respectively. **c,** Number of differentially expressed genes in each cell type between young and aged (left), and PD and aged (right). **d,** Circle plots showing differentially expressed example genes between young and aged midbrain. **e,** Heatmap representing significant GO terms and KEGG pathways in differentially expressed genes between young (left) and aged (right). **f,g,** As in **d,e**, for aged and PD midbrain.

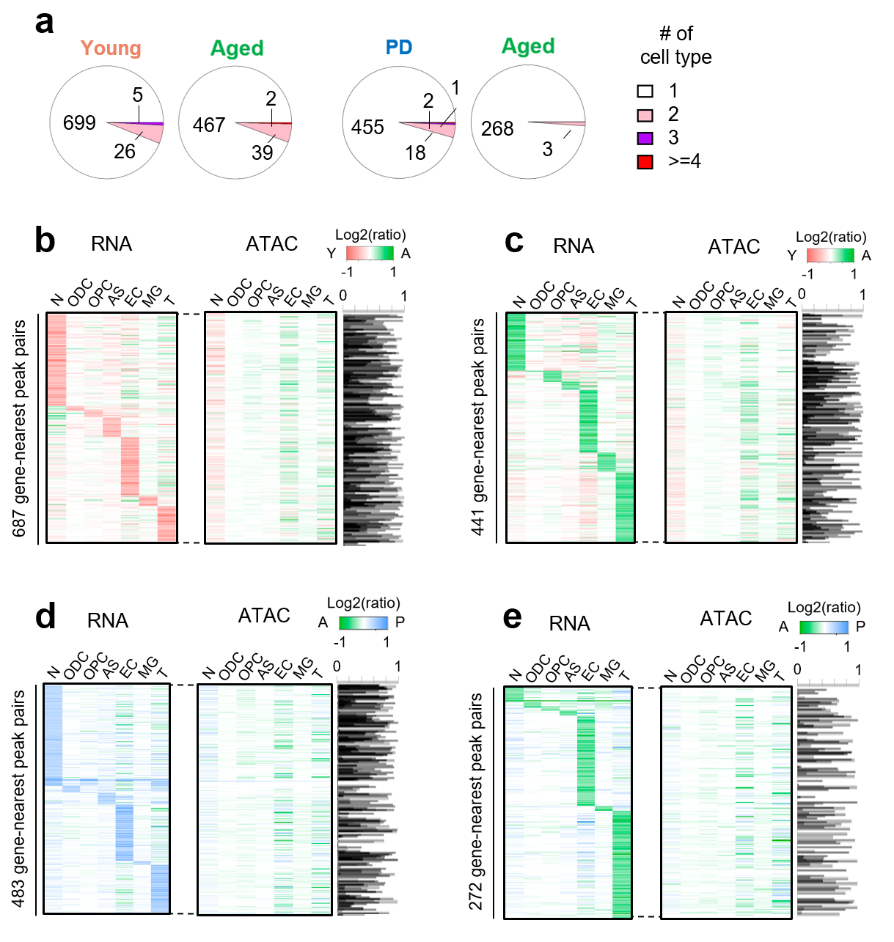

**Supplementary Fig. 3 | Cell-type specific DEG show low differential promoter activation.** **a,** Pie chart showing the number of differentially expressed genes that are commonly detected in multiple cell types. **b,c,** Heatmap showing RNA expression and ATAC intensity of young **(b)-** and aged **(c)-**enriched genes and their nearest peaks, respectively. Pearson correlation across cell types in each gene-peak pair is shown in the right Y-axis (**b,** Cor=0.319±0.342; **c,** 0.0952-2±0.536). **d-e,** Heatmap showing RNA expression and ATAC intensity of PD **(d)-** and aged **(e)-** enriched genes and their nearest peaks, respectively. Pearson correlation across cell types in each gene-peak pair is shown in the right Y-axis (**d,** Cor= 0.132±0.580; **e,** 0.0690±0.580).

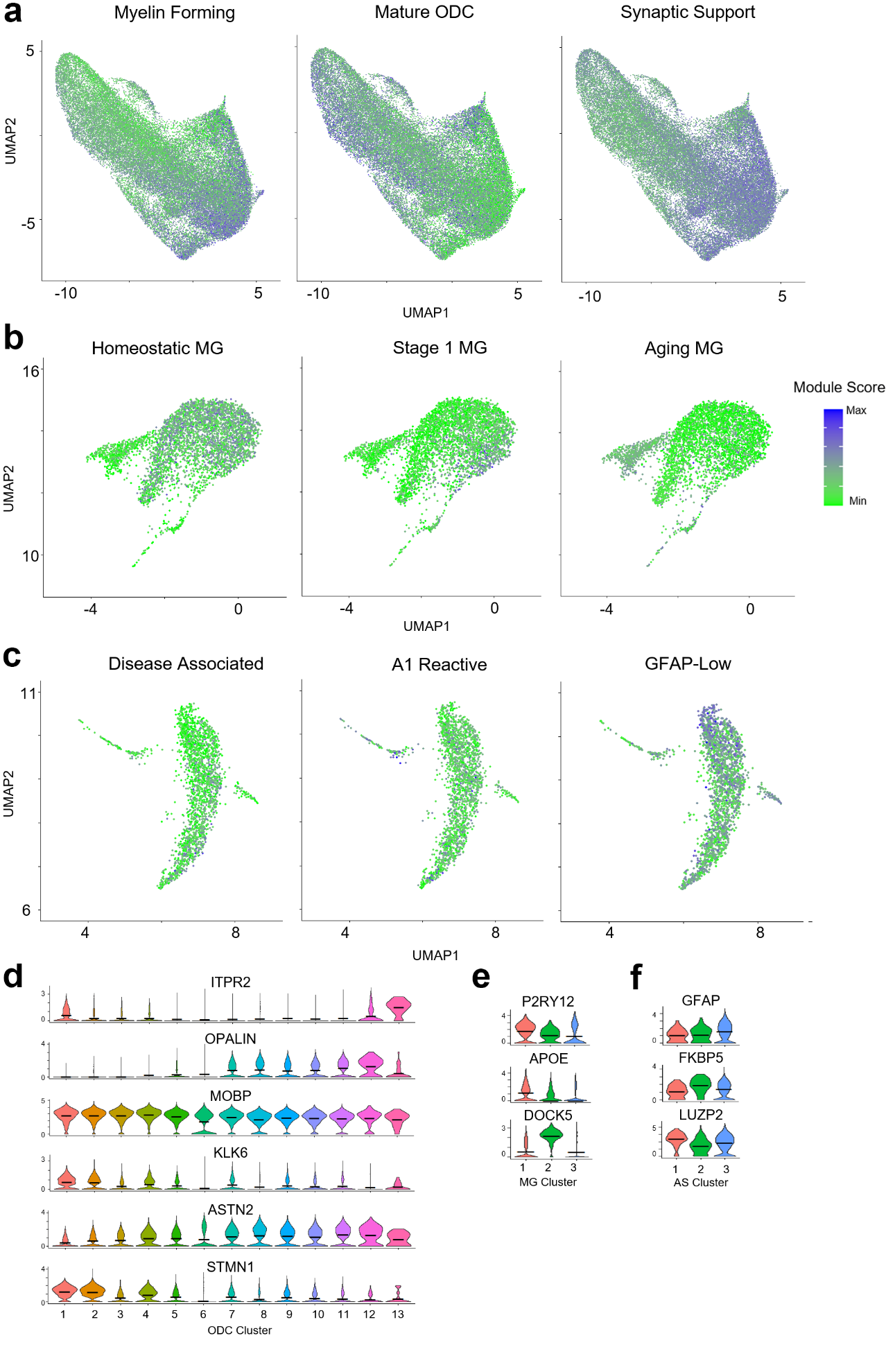

**Supplementary Fig. 4** **| Distinct subtype clusters**. **a-c,** UMAP of gene expression modules for previously identified functional cellular subtypes in the midbrain for ODC **(a)**, MG **(b)** and AS **(c)** show that distinct clusters of nuclei are enriched for different gene modules. **d-f,** Violin plots of expression for example genes from each of the modules used in ODC **(d)**, MG **(e)** and AS **(f)**.

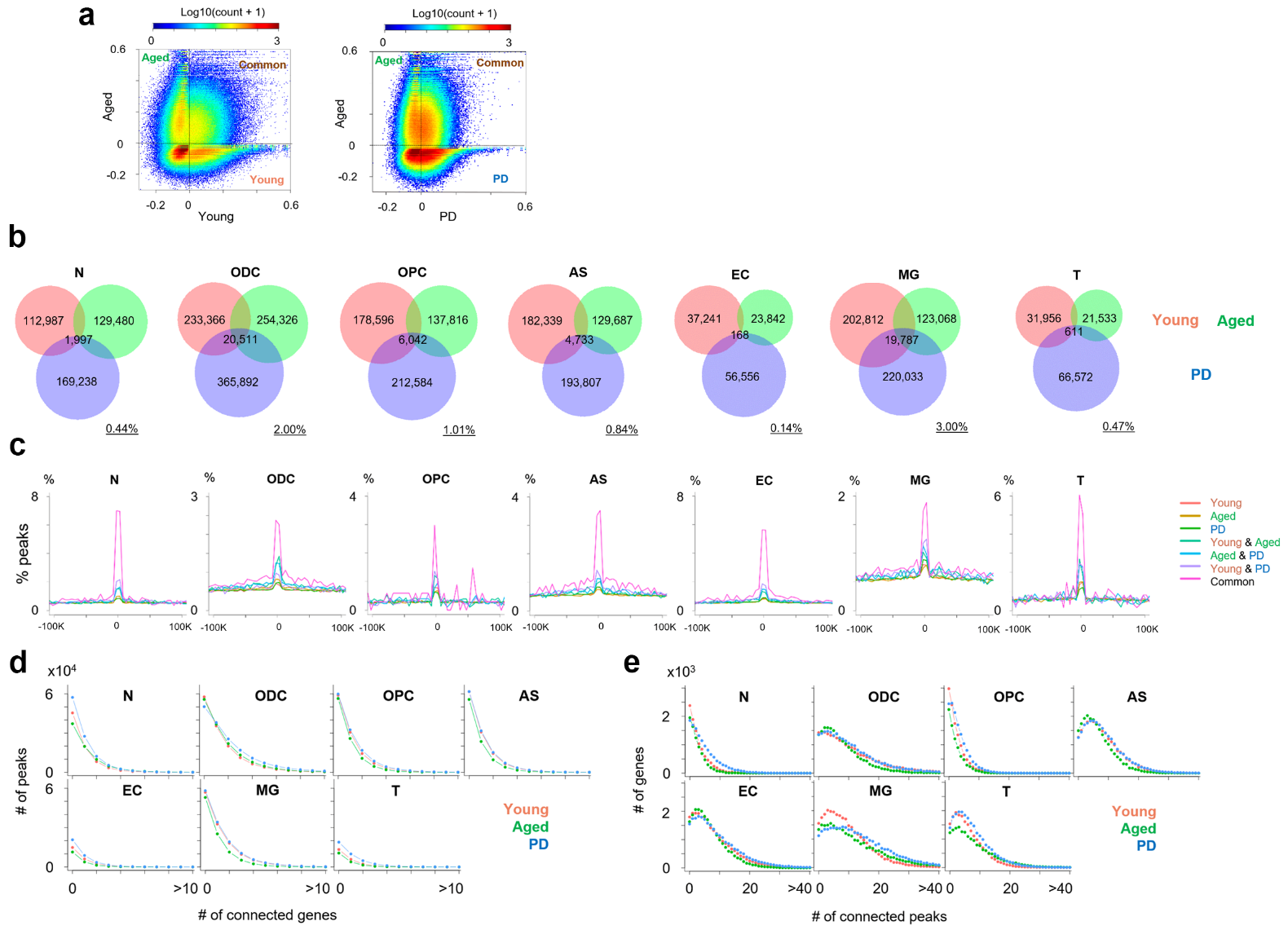

**Supplementary Fig. 5** **| Gene-peak connections are not well-conserved between groups.** **a,** Comparison of correlations of all gene-peak pairs (3,670,784 pairs) between young and aged nuclei (left), and aged and PD nuclei (right). Dashed line represents zero correlation coefficient. **b,** Venn diagram comparing significant gene-peak connections between young, aged and PD in each cell type. **c,** Graph showing distance of the connected peaks from TSSs between young, aged and PD. **d,** Histogram showing the number of connected genes per peak: Average 2.40 (young), 2.58 (aged) and 3.1 (PD) connected genes per peak. **e,** Histogram showing the number of connected peaks per gene: Average 10.6 (young), 13.4 (aged) and 15.1 (PD) connected peaks per gene.

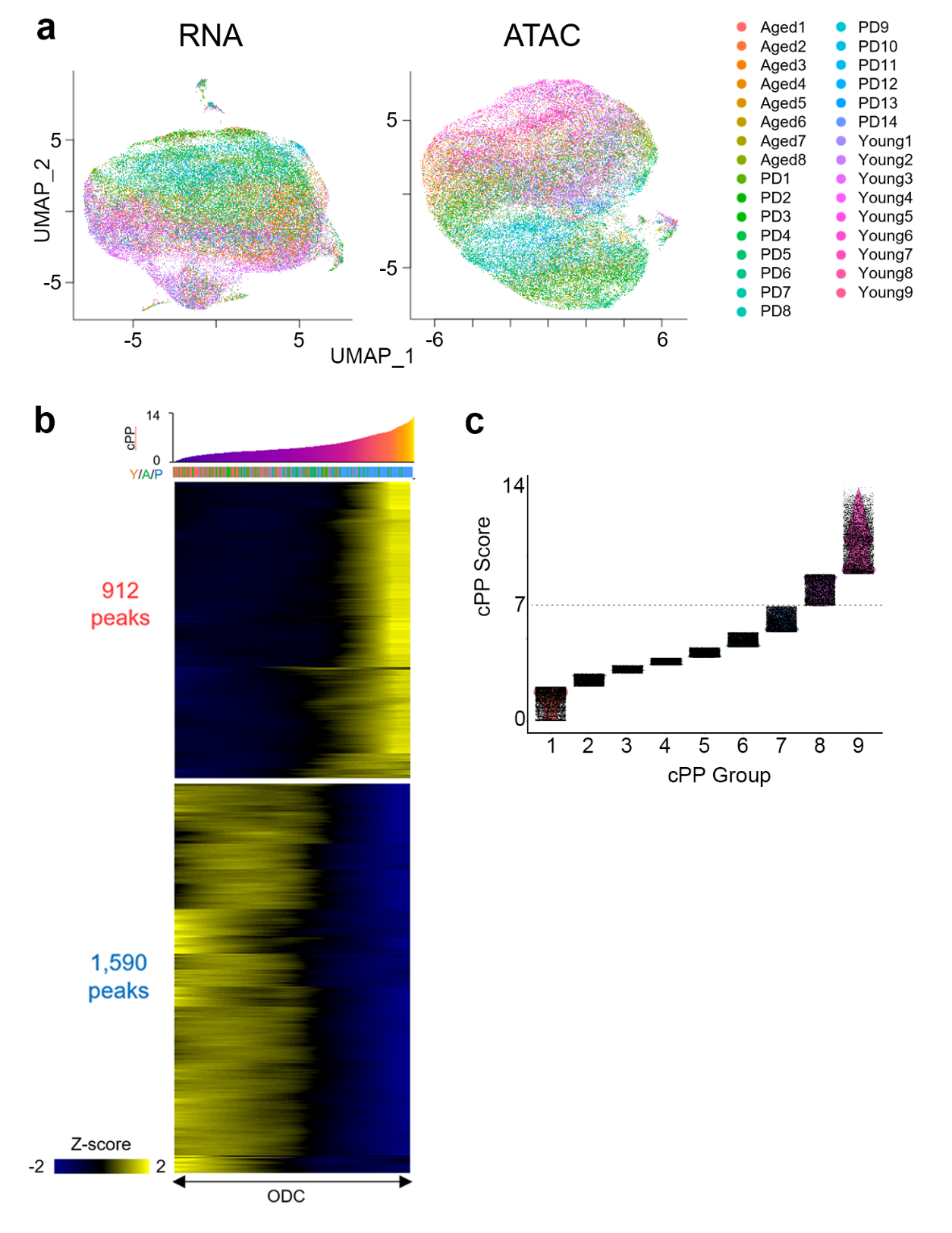

**Supplementary Fig. 6** **| Oligodendrocyte cPP.** **a,** UMAP plot of ODC nuclei colored by individual donors for RNA (left) and ATAC (right). **b,** Heatmap showing accessible ATAC peaks correlated with cPP trajectory. X-axis represents individual cells sorted by cPPs. Y-axis represents positively (upper)- and negatively (bottom)-correlated peaks (Cor > 0.1 or < -0.1). **c,** Plot of ODC nuclei divided into nine equal-sized groups based on cPP scores. Y-axis shows cPP score range in each group. Dotted line shows middle cPP score of 7.

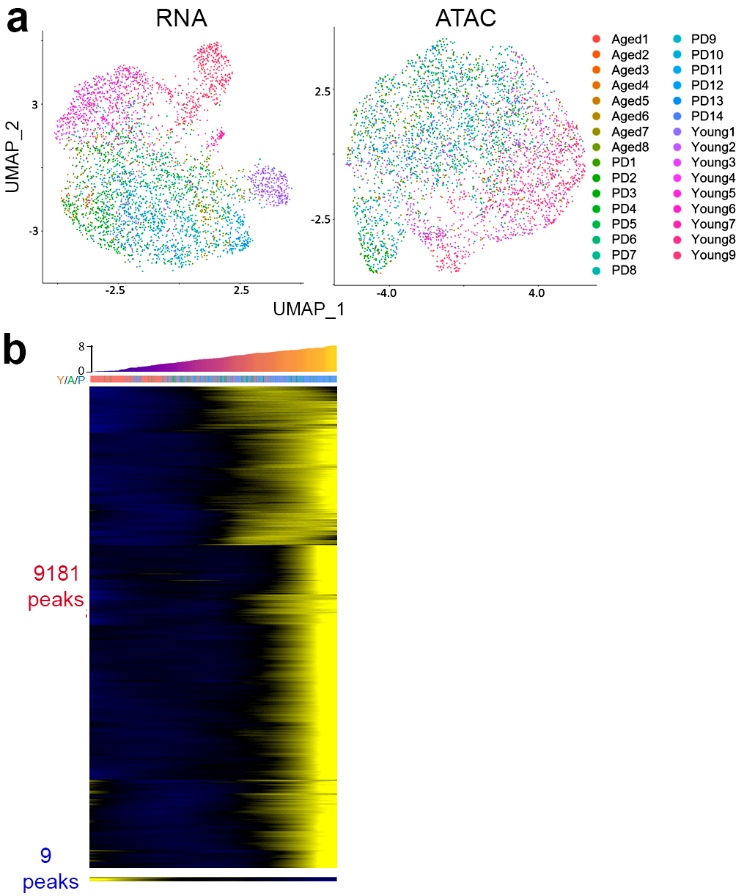

**Supplementary Fig. 7** **| Microglia cPP.** **a,** UMAP plot of MG nuclei colored by individual donors for RNA (left) and ATAC (right). **b,** Heatmap showing accessible ATAC peaks correlated with cPP trajectory. Y-axis of heatmap represents positively-(upper) and negatively(bottom)-correlated peaks (Cor > 0.1 or < -0.1).

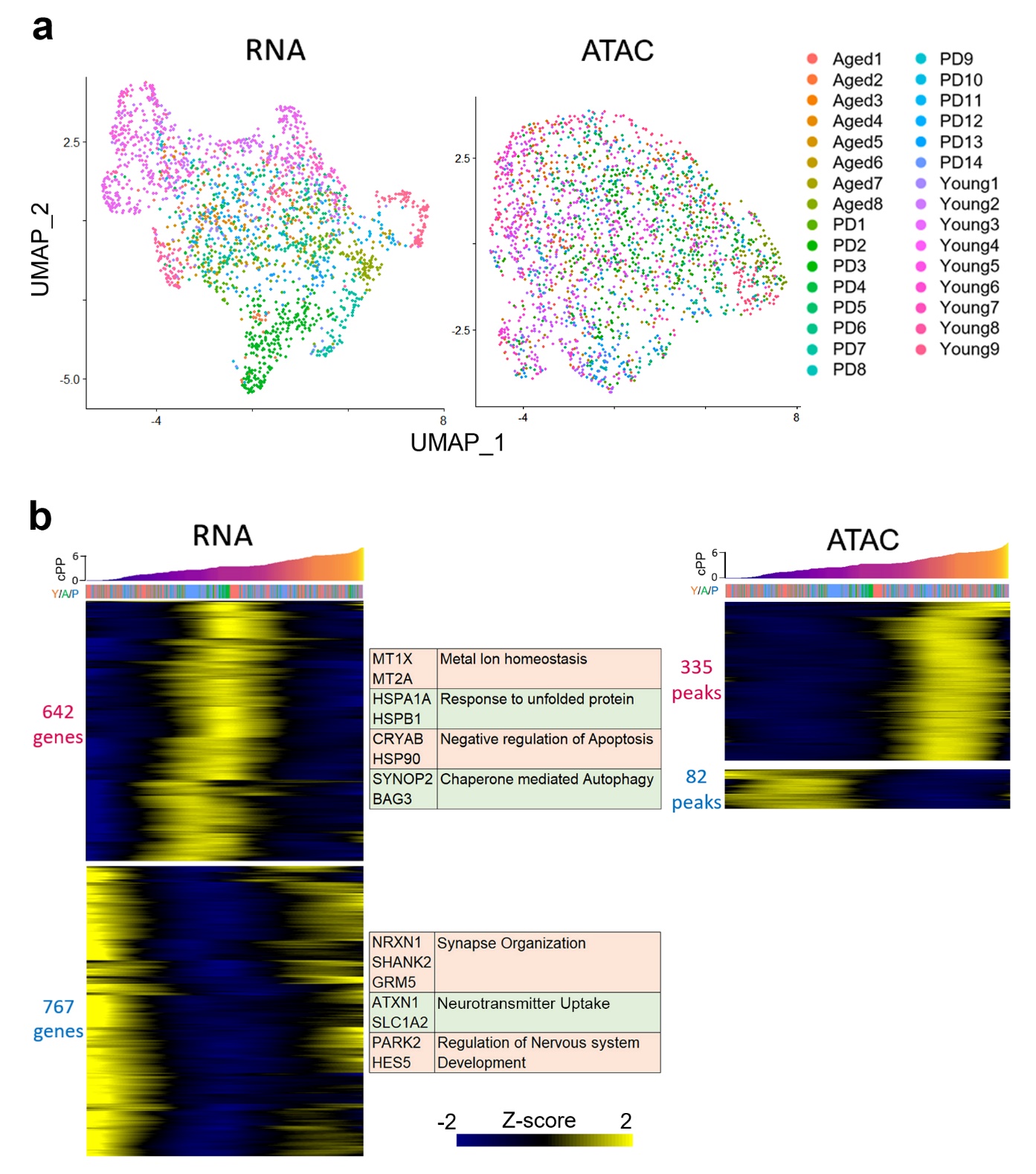

**Supplementary Fig. 8** **| Astrocyte cPP and DEG.** **a,** UMAP plot of AS nuclei colored by individual donors for RNA (left) and ATAC (right). **b,** Heatmap showing genes correlated with cPP trajectory (right). X-axis represents individual cells sorted by cPP. Y-axis of heatmap represents positively (upper)- and negatively (bottom)-correlated genes. Representative genes and significant GO terms are shown in the right panel (Cor > 0.1 or < -0.1). Heatmap showing accessible ATAC peaks correlated with cPP trajectory (left).

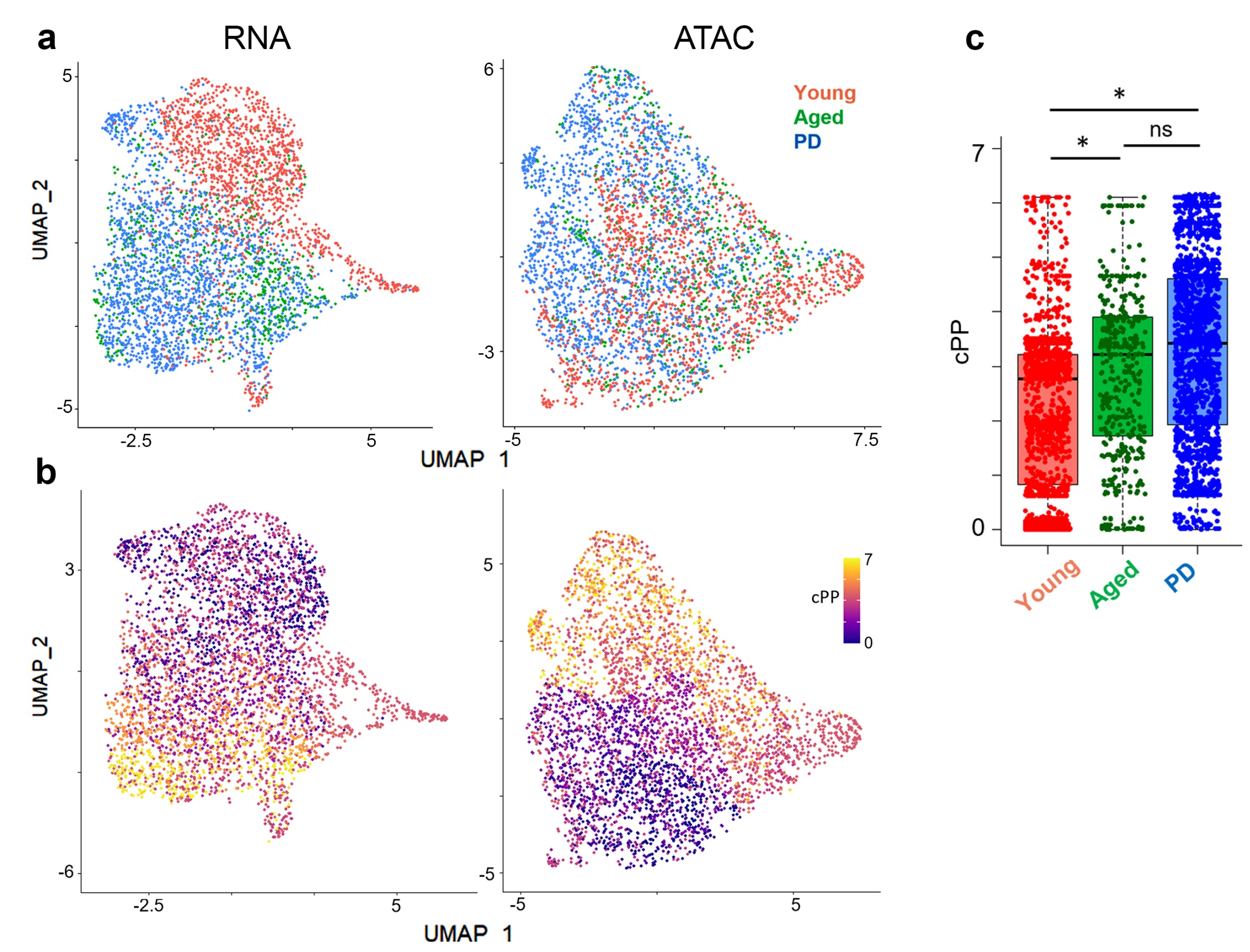

**Supplementary Fig. 9 | Establishment of pseudopathogenesis trajectory in OPC.** **a,b,** UMAP plot of OPC nuclei colored by young, aged and PD donor **(a)** or colored by cPP **(b)**. **c,** cPP scores of individual OPC nuclei from young, aged and PD midbrain are significantly changed over aging but not disease state. (One-way ANOVA, p-value for Y/A = 3.8e-63, p-value for A/P = 0.22).

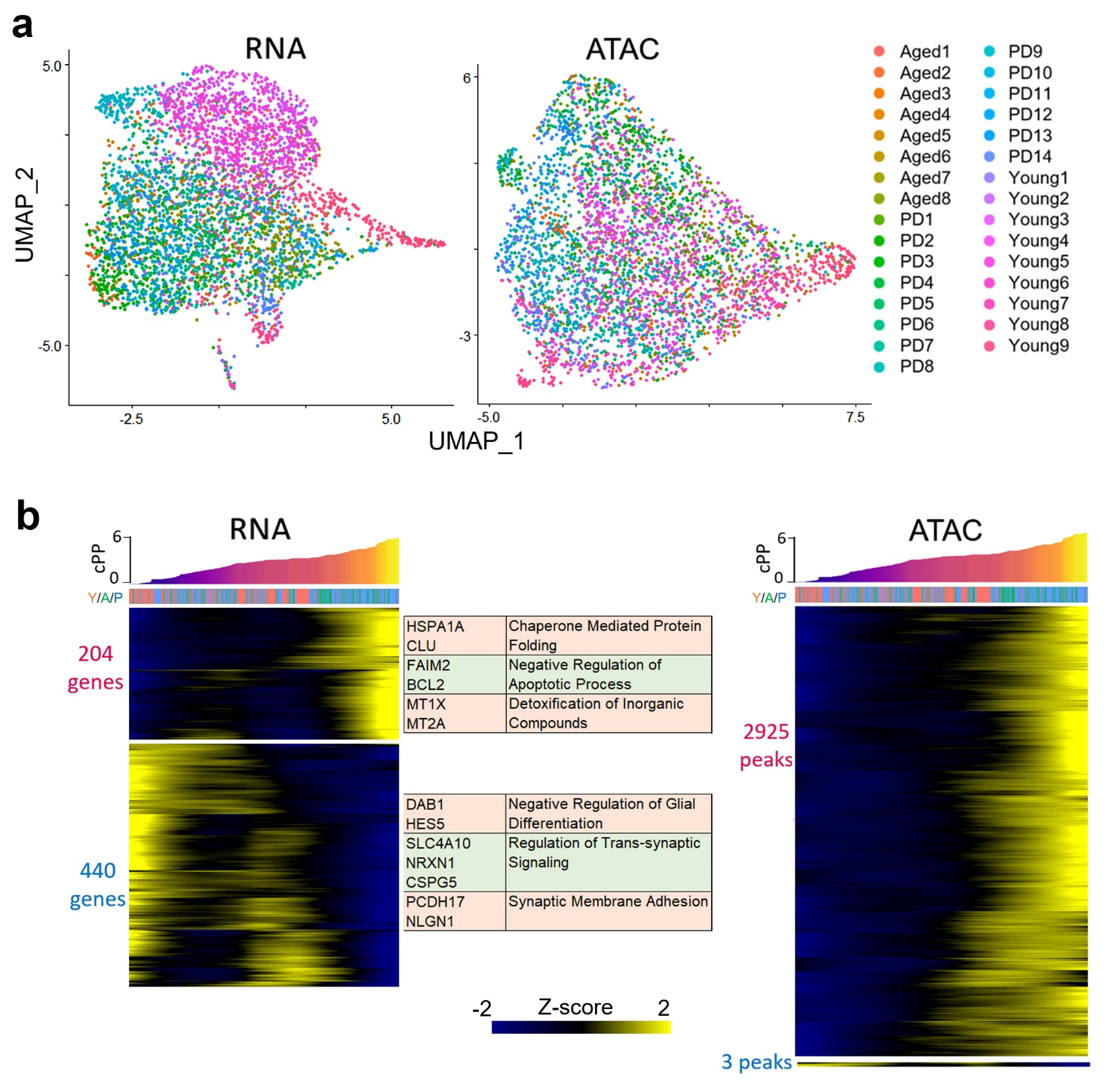

**Supplementary Fig. 10 | Oligodendrocyte Precursor Cell cPP and DEG.** **a,** UMAP plot of OPC nuclei colored by individual donors for RNA (left) and ATAC (right). **b,** Heatmap showing genes correlated with cPP trajectory (right). X-axis represents individual cells sorted by cPPs. Y-axis of heatmap represents positively (upper)- and negatively (bottom)-correlated genes. Representative genes and significant GO terms are shown in right panel (Cor > 0.1 or < -0.1). Heatmap showing accessible ATAC peaks correlated with cPP trajectory (left).

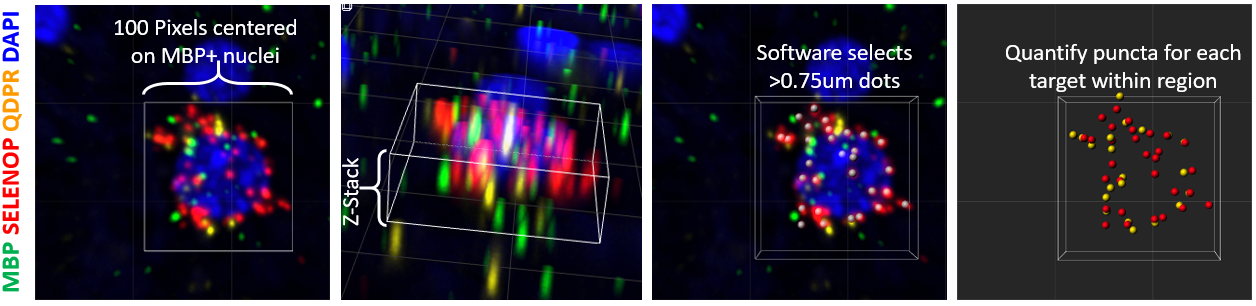

**Supplementary Fig. 11 | RNA-FISH quantification.** Using Imaris v9.9 imaging suite, we used the ‘Dots’ function for non-biased RNAScope puncta quantification of our confocal images. We selected a 100 pixel (18µm) square centered on MBP+ nuclei and quantified all puncta within the region on the stacked Z-plane (5µm-thickness tissue section).

**References for Supplementary Fig. 2a,b**

| Gene | Aging | PD | HD | AD | EPI | DD | ASD | BP | SCZ |
| --- | --- | --- | --- | --- | --- | --- | --- | --- | --- |
| NEAT1 | 1 | 2 | 3 | 4 | 5 |  |  |  | 6 |
| FKBP5 | 7 |  | 8 | 7 |  | 9 | 10 |  | 11 |
| NEGR1 |  |  |  |  |  | 12 |  | 13 | 14 |
| NRXN1 | 15 |  |  |  |  |  |  |  |  |
| KCTD8 |  |  |  |  |  |  |  |  |  |
| GRID2 |  |  |  | 16 |  |  |  |  |  |
| EML4 |  |  |  |  |  |  |  |  |  |
| Gene | Aging | PD | HD | AD | EPI | DD | ASD | BP | SCZ |
| SEM1 |  |  |  |  |  |  |  |  |  |
| SLC38A2 |  | 17 | 18 | 19 |  |  |  |  |  |
| HSP90AA1 | 20 | 21 | 22 | 23,24 | 25 |  | 26 | 11 | 27 |
